# Widespread alterations of diffusion tensor imaging metrics in patients with schizophrenia without current auditory hallucinations

**DOI:** 10.1101/2023.04.18.23288743

**Authors:** Stener Nerland, Nora Berz Slapø, Claudia Barth, Lynn Mørch-Johnsen, Kjetil Nordbø Jørgensen, Dani Beck, Laura A. Wortinger, Lars T. Westlye, Erik G. Jönsson, Ole A. Andreassen, Ivan I. Maximov, Oliver M. Geier, Ingrid Agartz

**Author notes:** <u>Corresponding author:</u> Stener Nerland.

## Abstract

**Background:** Studies have linked auditory hallucinations (AH) in schizophrenia-spectrum disorders (SCZ) to altered cerebral white matter microstructure within the language and auditory processing circuitry (LAPC). However, the neuroanatomical distribution and specificity to the LAPC remains unclear. Here, we investigated the relationship between AH and DTI among patients with SCZ using diffusion tensor imaging (DTI).

**Methods:** We included patients with SCZ with (AH+; n=59) and without (AH-; n=81) current AH, and 140 age-and-sex-matched controls. Fractional anisotropy (FA), mean diffusivity (MD), radial diffusivity (RD), and axial diffusivity (AD) were extracted from 39 fibre tracts. We used principal component analysis (PCA) to identify general factors of variation across fibre tracts and DTI metrics. Regression models adjusted for sex, age, and age^2^ were used to compare tract-wise DTI metrics and PCA factors between AH+, AH-, and healthy controls and to assess associations with clinical characteristics.

**Results:** Widespread differences relative to controls were observed for MD and RD in patients without current AH. Only limited differences in two fibre tracts were observed between AH+ and controls. Unimodal PCA factors based on MD, RD, and AD, as well as multimodal PCA factors, differed significantly relative to controls for AH-, but not AH+. We did not find any significant associations between PCA factors and clinical characteristics.

**Conclusions:** Contrary to previous studies, DTI metrics differed mainly in patients *without* current AH compared to controls, indicating a widespread neuroanatomical distribution. Our results challenge the notion that altered DTI metrics in the LAPC is a specific feature underlying AH.

## 1 Introduction

Auditory hallucinations (AH), i.e., auditory percepts not elicited by an external source, is a core symptom in schizophrenia spectrum disorders (SCZ). AH has a lifetime prevalence of 60-80% in SCZ (Waters et al., 2014) and can significantly affect quality of life (Chiang, Beckstead, Lo, & Yang, 2018; Choi et al., 2021). Although the pathophysiology underlying AH is poorly understood, magnetic resonance imaging (MRI) studies have implicated alterations in cerebral white matter (WM) microstructure (Lee et al., 2009; Seok et al., 2007; Shao, Liao, Gu, Chen, & Tang, 2021; Shergill et al., 2007), regional morphology of the cerebral cortex (Mørch-Johnsen et al., 2017; Neckelmann et al., 2006), and functional activation during active hallucination (Kompus, Westerhausen, & Hugdahl, 2011). Notably, many studies have reported alterations in the language and auditory processing circuitry (LAPC; Friederici & Gierhan, 2013; Hickok, 2012) in patients with SCZ and AH, including cortical regions such as Heschl’s gyrus (Mørch-Johnsen et al., 2017), Broca’s area (Fovet et al., 2022), and Wernicke’s area (Salisbury, Wang, Yeh, & Coffman, 2021), as well as the WM fibre tracts connecting them. In particular, it has been hypothesised that disrupted connectivity within the LAPC results in erroneous source attribution of voices, which in turns leads to verbal AH (Curcic-Blake et al., 2015; Feinberg, 1978; Feinberg & Guazzelli, 1999; Frith, 2005).

Diffusion tensor imaging (DTI) studies have reported alterations in WM fibre tracts within the LAPC in patients with SCZ and AH, including group differences between patients with and without AH and associations with AH severity (Hubl et al., 2004; Psomiades et al., 2016; Sato et al., 2021; Zhang et al., 2018). Given its crucial role in language processing, the left arcuate fasciculus (AF), a heavily myelinated fibre bundle that connects Broca’s and Wernicke’s areas, has been frequently studied in this context (Friederici & Gierhan, 2013; Wernicke, 1874). However, results have been inconsistent, with reports of both higher (Chawla, Deep, Khandelwal, & Garg, 2019; Hubl et al., 2004; Psomiades et al., 2016) and lower (Curcic-Blake et al., 2015; McCarthy-Jones, Oestreich, & Whitford, 2015; Oestreich, McCarthy-Jones, Whitford, & Australian Schizophrenia Research Bank, 2016) fractional anisotropy (FA) in patients with AH. Using tract-based spatial statistics (TBSS), Curcic-Blake et al. (2015) found lower FA in SCZ with verbal AH compared to controls within additional fibre tracts, including the left inferior fronto-occipital fasciculus (IFO), the left uncinate fasciculus (UF), and the right superior longitudinal fasciculus (SLF). Studies have also implicated interhemispheric auditory pathways (Curcic-Blake et al., 2015; Knöchel et al., 2012; Mulert et al., 2012; Wigand et al., 2015). Since most previous studies have focused on a limited number of fibre tracts, the full extent of the relationship between AH in SCZ and WM microstructure remains unclear.

Associations with DTI metrics are often ascribed to microstructural tissue properties such as degree of myelination, fibre tract organisation, axonal ordering and density, and membrane permeability. However, the specificity is low and DTI metrics reflect a combination of neurobiological processes (Beaulieu, 2002; Jelescu, Palombo, Bagnato, & Schilling, 2020). In particular, the interpretation of FA as a measure of white matter integrity has been questioned (Jones, Knösche, & Turner, 2013). While it is not possible to directly link DTI metric alterations to microstructure, DTI metrics exhibit different sensitivities to distinct tissue properties. For instance, joint diffusion and histological studies in the cuprizone model of demyelination in mice indicate that radial diffusivity (RD) is related to de- and re-myelination (Guglielmetti et al., 2016; Song et al., 2005; Sun et al., 2006), whereas axonal diffusivity (AD) may be more sensitive to axonal damage (Winklewski et al., 2018). Similarly, mean diffusivity (MD) has been reported to be more sensitive to myelin-staining indices than either FA or RD (Seehaus et al., 2015). Analysing a wider range of DTI metrics could therefore be useful for investigating putative WM microstructural correlates of AH in SCZ.

Leveraging the shared variation across DTI metrics and fibre tracts may provide additional information beyond analysing each metric and fibre tract in isolation (Cox et al., 2016; Geeraert, Chamberland, Lebel, & Lebel, 2020; Vaher et al., 2022). For example, Chamberland et al. (2019) used principal component analysis to identify components that explained 80% of the variation across fibre tracts and diffusion measures. Their results revealed age-related effects, some of which were not detectable at the level of individual DTI metrics. Importantly, the components loaded onto diffusion measures with shared sensitivities to specific tissue properties, demonstrating that this approach yields biologically interpretable information. Using a similar approach, Vaher et al. (2022) found evidence for generalised dysmaturation in individuals who were born preterm. Interestingly, they found that while the effect of gestational age on diffusion measures was mostly shared, there was an additional independent effect on specific fibre tracts. Thus, combining DTI metrics can enhance sensitivity to group differences and relevant brain-behaviour associations (Alnæs et al., 2018; Alnæs, Kaufmann, Marquand, Smith, & Westlye, 2020; Tønnesen et al., 2020).

In the present study, we investigated the relationship between current AH, i.e., within a week of clinical inclusion, and FA, MD, RD, and AD in a well-powered sample of patients with SCZ and age-and-sex-matched healthy controls. We included a broad range of fibre tracts and DTI metrics and assessed variation across both fibre tracts and DTI metrics using an established dimensionality reduction framework (Chamberland et al., 2019; Geeraert et al., 2020; Vaher et al., 2022). Based on previous studies, we hypothesised lower FA and higher MD and RD in fibre tracts within the LAPC in patients with current AH (Curcic-Blake et al., 2015; Sato et al., 2021; Shao et al., 2021). Given the paucity of studies on links between AH and AD, these analyses were exploratory. Follow-up analyses on lifetime AH were performed to examine vulnerability towards experiencing AH as a trait marker. We assessed the effects of psychotic symptom severity, age at onset, duration of illness, and antipsychotic medication use and dose in exploratory analyses. Finally, since both head size and sex contributes to variation in DTI metrics (Eikenes, Visser, Vangberg, & Håberg, 2023), we assessed the effect of sex and intracranial volume (ICV) on putative group differences.

## 2 Methods

### 2.1 Participants

Participants diagnosed with SCZ and age-and-sex-matched healthy controls were included from the ongoing Thematically Organised Psychosis (TOP; n=659) study at the Oslo University Hospital, Norway, and from the Human Brain Informatics (HUBIN; n=92) project at Karolinska Institutet in Stockholm, Sweden. Patients were referred from psychiatric departments and outpatient clinics in the greater Oslo region and from catchment areas within the North-Western Stockholm County respectively. Healthy controls were recruited based on population registries for both TOP and HUBIN and among hospital staff for HUBIN.

Exclusion criteria included an age outside the range 18-65 years, intelligence quotient (IQ) less than 70, and neurologic illness or previous moderate to severe head injury. Controls were also excluded if they had a history of substance abuse/dependency or a first-degree relative diagnosed with a severe psychiatric disorder.

Healthy controls were age-and-sex-matched to the patient group using genetic matching without replacement in the *MatchIt* package (Ho, Imai, King, & Stuart, 2011) in R. We used the model Diagnosis ∼ Age + Sex + Scanner with exact matching on MRI scanner. This approach avoids issues with propensity score matching, which does not ensure close pairings of participants (King & Nielsen, 2019).

The final study sample (n=280) included 140 age-and-sex-matched healthy controls (mean age: 33.2; range=[18.4, 63.6]; 35.7% female and 140 patients with SCZ (mean age: 33.3; range=[18.5, 63.6]; 35.7% female) diagnosed with schizophrenia (n=106), schizophreniform disorder (n=24), and schizoaffective disorder (n=10).

### 2.2 Clinical assessment

For patients in TOP, diagnoses and lifetime symptoms were assessed using the Structured Clinical Interview for DSM-IV axis 1 disorders (SCID-IV; Spitzer, Williams, Gibbon, & First, 1992), and current symptoms, i.e., the week before clinical assessment, were evaluated with the Positive and Negative Syndrome Scale, PANSS (Kay, Fiszbein, & Opler, 1987). PANSS scores were converted to symptom factors from the Wallwork five-factor model (Wallwork, Fortgang, Hashimoto, Weinberger, & Dickinson, 2012), i.e., positive (P1 + P3 + P5 + G9), negative (N1 + N2 + N3 + N4 + N6 + G7), disorganised (P2 + N5 + G11), excited (P4 + P7 + G8 + G14), and depressive (G2 + G3 + G6) symptom factors. For patients in HUBIN, diagnoses and lifetime symptoms were assessed using the SCID-III-R (Spitzer et al., 1992) and Schedules for Clinical Assessment in Neuropsychiatry (SCAN; Wing et al., 1990), and current symptoms were evaluated with the Scales for the Assessment of Positive and Negative Symptoms, SAPS and SANS (Andreasen, 1983, 1984).

Antipsychotic medication use was determined via interviews. To compare doses across antipsychotic medication type and dosage, we converted antipsychotic medication dosage to chlorpromazine (CPZ; Jørgensen et al., 2016) equivalent doses (mg/day). We defined age at onset as age at first psychotic episode (verified by SCID-IV, SCID-III-R, or SCAN) and duration of illness as years from age at onset to age at MRI scan. IQ was assessed with the Wechsler Abbreviated Scale of Intelligence (WASI-II) in TOP and the Wechsler Adult Intelligence Scale (WAIS) in HUBIN. Alcohol and drug use was assessed with the Alcohol Use Disorder Identification Test (AUDIT; Saunders, Aasland, Babor, De La Fuente, & Grant, 1993) and the Drug Use Disorders Identification Test (DUDIT; Berman, Bergman, Palmstierna, & Schlyter, 2005). Psychosocial functioning was rated using the split version of the Global Assessment of Functioning Scale (GAF; Pedersen, Hagtvet, & Karterud, 2007).

We determined current AH status based on the PANSS-P3 item in TOP and the SAPS-H1 item in HUBIN. For TOP, we defined the current AH group (AH+) as those with a P3 score ≥ 3, and the current non-AH group (AH-) as those with a P3 score < 3. For HUBIN, we defined the current AH group as having a H1 score ≥ 2, and the current non-AH group (AH-) as H1 < 2. Presence (L-AH+) or absence (L-AH-) of lifetime AH was determined with the SCID-B16 item for TOP, with L-AH-defined as subthreshold/not present. For HUBIN, lifetime AH was determined with the 17.004 item in SCAN, with L-AH-defined as absent/not clinically significant or mild/questionable. See **Supplementary Note 1** for additional information on clinical assessment of AH.

### 2.3 MRI acquisition and processing

MRI data was acquired on three different 3T scanner platforms; a GE Signa HDxt and a GE Discovery MR750 at Oslo University Hospital (OUS), Ullevål, Oslo, and a GE Discovery MR750 at Karolinska Institutet in Stockholm, Sweden. See **Supplementary Table 3** for acquisition parameters. DTI images were processed using an optimised pipeline (Maximov, Alnæs, & Westlye, 2019) and FSL (Jenkinson, Beckmann, Behrens, Woolrich, & Smith, 2012; version 6.0.3) with corrections for noise (Veraart et al., 2016), Gibbs ringing (Kellner, Dhital, Kiselev, & Reisert, 2016), echo-planar imaging (EPI) motion, eddy currents, and susceptibility distortions. DTI metrics were estimated with *dtifit* using the linear weighted least squares algorithm.

Fibre tractography was performed with XTRACT (Warrington et al., 2020) in FSL using well-validated and robust protocols for automated fibre tractography. Median FA, MD, RD, and AD were extracted for 39 inter- and intra-hemispheric fibre tracts. See **Figure 1** and **Table 2** for overviews of the included fibre tracts and **Supplementary Note 2** for information on fibre tract selection. To compute ICV, we used Sequence Adaptive Multimodal Segmentation (SAMSEG; Puonti, Iglesias, & Van Leemput, 2016) with default parameters using a single T1-weighted image as input.

**Table 1.**
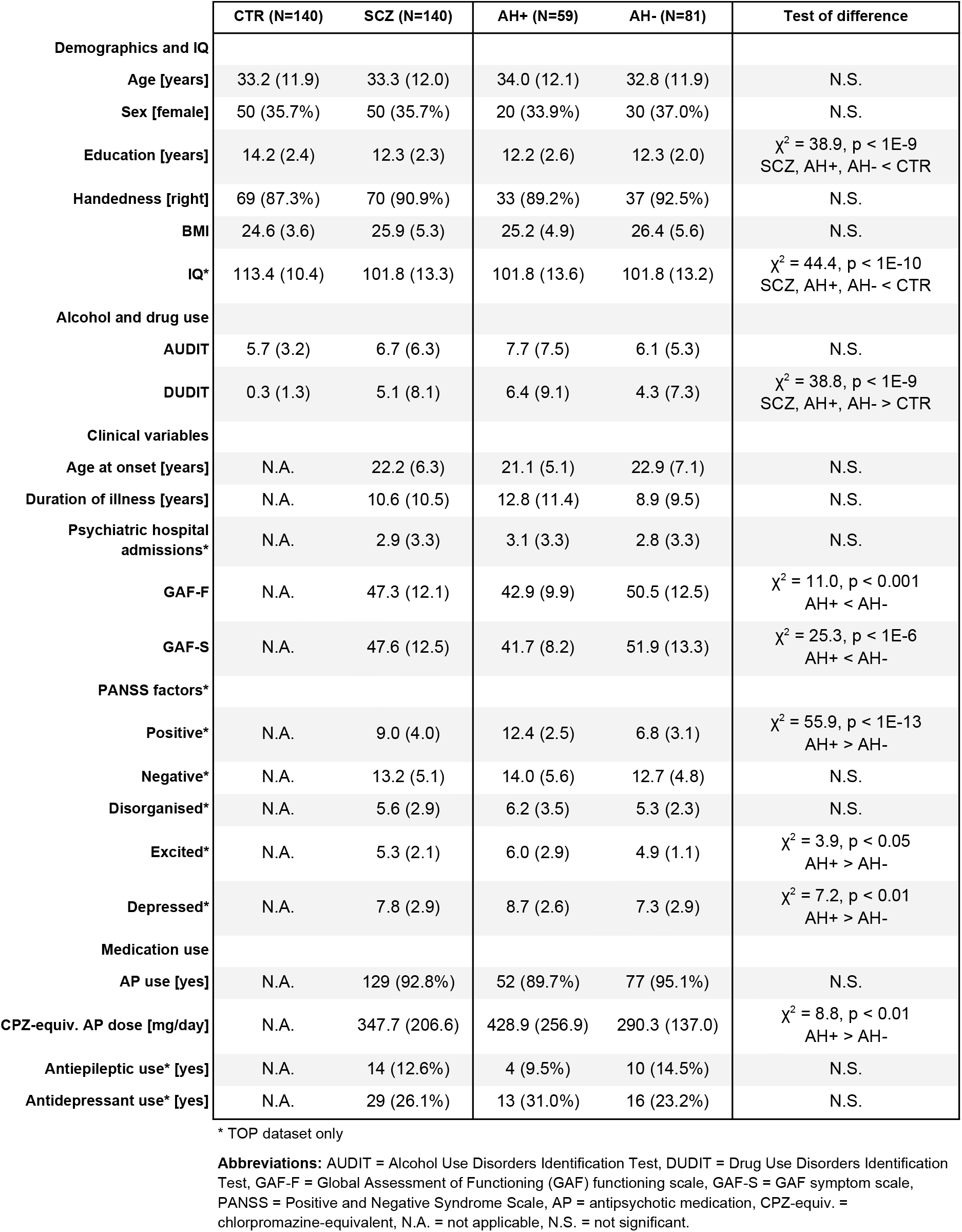
Demographic and clinical characteristics of the healthy controls, patients, and patient subgroups with (AH+) and without (AH-) current hallucinations. Continuous variables are reported as mean (standard deviation) and categorical variables as count (percentage).

**Table 2.**
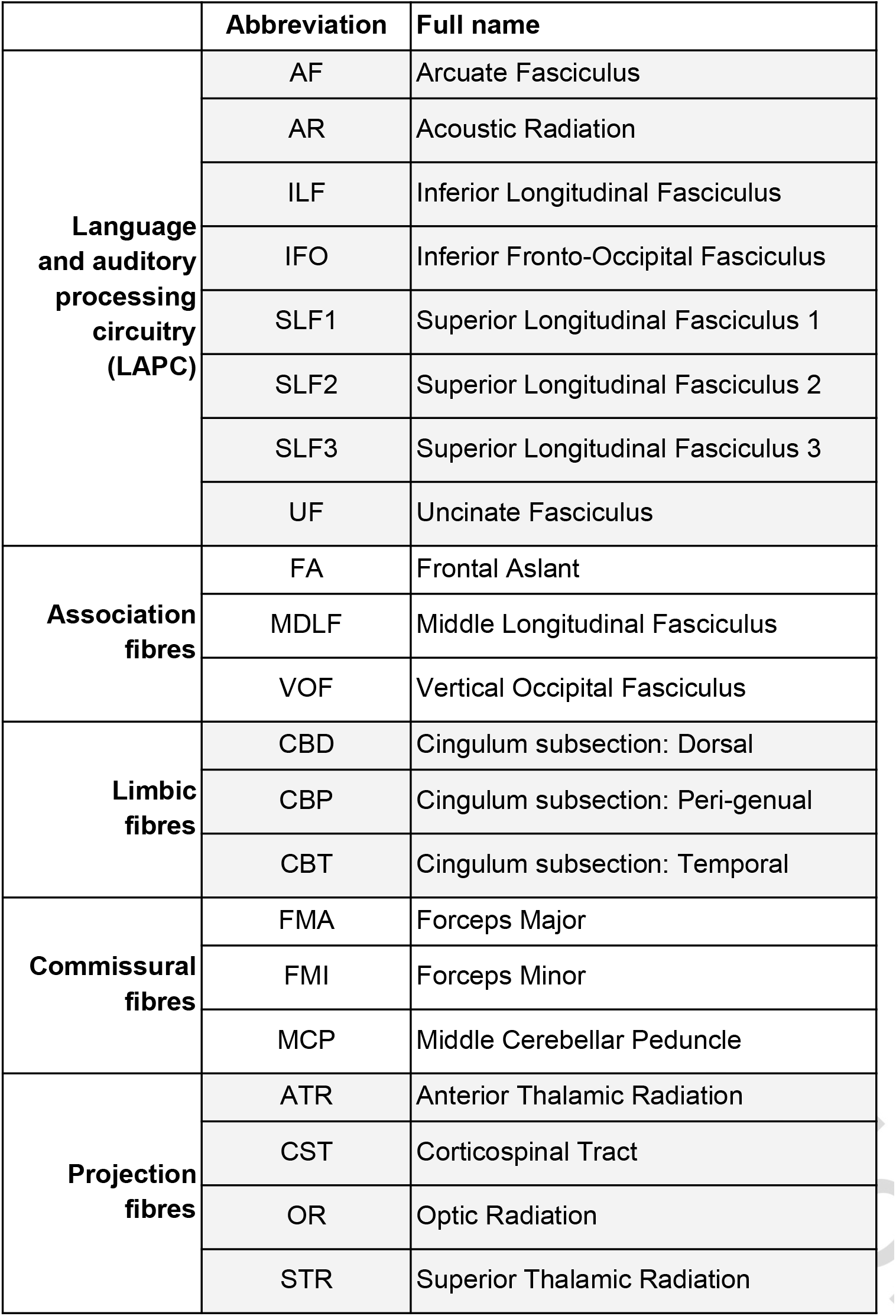
Overview of the 39 fibre tracts included in the study. With the exception of the commissural fibre tracts, FMA, FMI, and MCP, the tracts are divided into left and right hemispheres.

**Figure 1.**
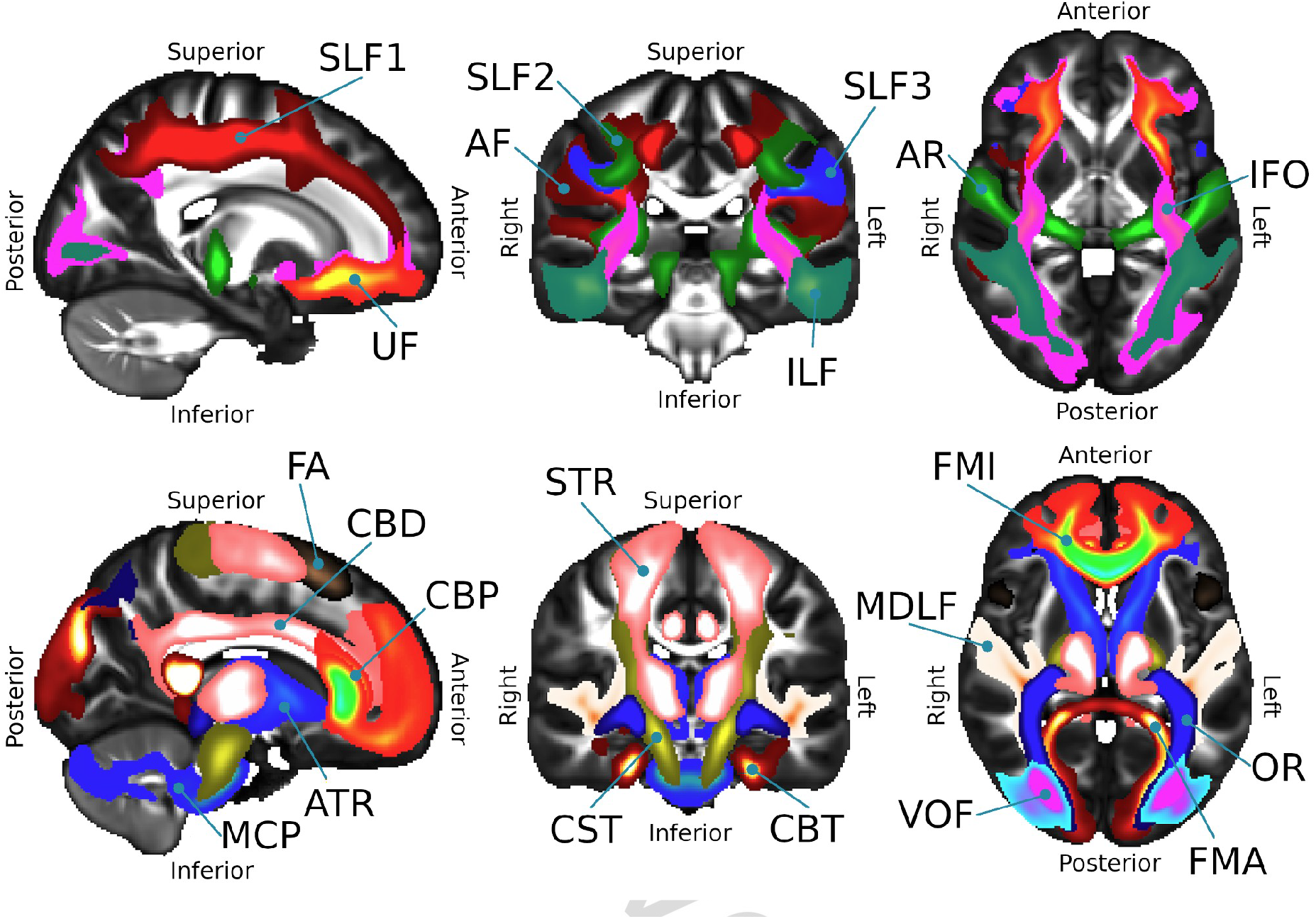
Visualisation of the fibre tracts included in the present study. The top row shows fibre tracts within the language and auditory processing circuitry (LAPC). The bottom row shows the other fibre tracts.

The batch-adjustment algorithm ComBat (Fortin et al., 2017) was used to adjust for non-biological variation in DTI metrics due to differences in pulse sequence and scanner platform. Age, sex, and diagnosis were entered as variables of interest. See **Supplementary Figure 1** for boxplots showing the effect of scanner harmonisation and **Supplementary Note 3** for information on quality assurance procedures.

### 2.4 Statistical analyses

Statistical analyses were performed in R (version 4.2.3; R Core Team, 2018). All regression models included age, age^2^, and sex as covariates, where we included age^2^ since the relationship between age and DTI metrics may be nonlinear (Beck et al., 2021). Cohen’s d effect sizes (d) were obtained from t-values using the *effectsize* package (Ben-Shachar, Lüdecke, & Makowski, 2020) implemented in R. We adjusted p-values for the false discovery rate (FDR) using the Benjamini-Hochberg procedure (Benjamini & Hochberg, 1995). An FDR-adjusted p-value < 0.05 was considered statistically significant.

#### 2.4.1 Demographic and clinical group comparisons

We assessed group differences in age, sex, years of education, handedness, BMI, IQ, AUDIT and DUDIT scores, age at onset, duration of illness, number of psychiatric hospital admissions, GAF functioning (GAF-F) and symptom (GAF-S) scores, the five Wallwork symptom factors, antipsychotic use (yes/no), CPZ-equivalent doses, and antiepileptic and antidepressant use. Continuous and categorical variables were compared using the Kruskal-Wallis test and the χ^2^ test respectively.

#### 2.4.2 Tract- and metric-specific analyses

To assess group differences in tract-specific DTI metrics in AH+ and AH-compared to healthy controls, we fitted univariate regression models with DTI metrics (FA, MD, RD, or AD) in each of the 39 fibre tracts as dependent variables and current AH status as the variable of interest. To assess tract-wise group differences between AH+ and AH-for each DTI metric, we fitted similar models among AH+ and AH-only.

In post hoc analyses, we explored group differences in tract-specific DTI metrics in patients with (L-AH+) and without (L-AH-) a lifetime history of auditory hallucinations compared to healthy controls and each other. Finally, we compared tract-specific DTI metrics in patients with SCZ, i.e., current AH+ and AH-combined, with those of controls in regression models where diagnostic group was the independent variable of interest.

#### 2.4.3 Uni- and multimodal general factor analyses

To assess the patterns of associations with AH status across WM fibre tracts and DTI metrics, we extracted uni- and multimodal general factors (g-factors) following a previously established dimensionality reduction framework (Chamberland et al., 2019; Geeraert et al., 2020; Vaher et al., 2022). This entailed conducting principal component analysis (PCA) across fibre tracts for each DTI metric to create unimodal g-factors and across both fibre tracts and DTI metrics to create multimodal g-factors. PCA was conducted by singular value decomposition on scaled tract-specific DTI metrics using the *prcomp* function in R. To assess the suitability of the data for factor analysis, we used the Kaiser-Meyer-Olkin test (Dziuban & Shirkey, 1974) and Bartlett’s test of sphericity, which measure the amount of shared variance and the overall degree of correlation within the variables.

Unimodal g-factors were created by conducting PCA on all 39 fibre tracts, for each DTI metric separately. Based on the literature (Chamberland et al., 2019; Geeraert et al., 2020; Vaher et al., 2022), we aimed to identify one general factor for each DTI metric to capture shared variation across fibre tracts. We therefore extracted the first principal components. This yielded four g-factors, g-FA, g-MD, g-RD, and g-AD, quantifying the proportion of shared variation across fibre tracts for each DTI metric.

Multimodal g-factors were created by conducting PCA on all tracts and DTI metrics simultaneously. That is, we performed PCA on a matrix with rows corresponding to subject-tract pairs (i.e., 39 rows per subject) and four columns corresponding to FA, MD, RD, and AD measurements. Prior studies found that two components were necessary to capture multimodal variation (Chamberland et al., 2019; Geeraert et al., 2020; Vaher et al., 2022). We therefore extracted the first and second principal components, resulting in two multimodal g-factors, g-Dim1 and g-Dim2, quantifying the proportion of shared variation across both DTI metrics and fibre tracts.

To compare uni- and multi-modal g-factors in AH+, AH- with those of healthy controls, we fitted separate regression models with current AH (AH+/AH-) as the variable of interest and each g-factor as dependent variables. We fitted similar models among patients to directly compare g-factors of AH+ with those of AH-.

#### 2.4.4 Associations with clinical and demographic variables

We performed additional analyses to investigate if clinical characteristics other than AH status were associated with DTI measurements. Among patients we assessed relationships between uni- and multimodal g-factors and the following clinical variables: Age at onset, duration of illness, antipsychotic medication use (yes/no), CPZ-equivalent dose, and the five Wallwork symptom factors (positive, negative, depressed, disorganised, and excited). In these analyses, we fitted separate univariate regression models for each clinical measure as variables of interest and each g-factor as dependent variables.

Next, we examined if group differences in g-factors were confounded by ICV or BMI. To do this, we fitted separate regression models with ICV and BMI included as independent variables together with the current AH term. We fitted the models both in the whole sample, for the contrast of AH+ and AH- with controls, and among patients only, for the direct comparison of AH+ with AH-.

Finally, we examined if group differences in current AH differed by age or sex. To do this, we fitted regression models including interaction terms for current AH by age and age^2^ and sex, as well as their respective main effects. Uni- and multimodal g-factors were specified as dependent variables and separate models were fitted for each putative interaction.

## 3 Results

### 3.1 Demographic and clinical group comparisons

Years of education and IQ were lower in patients, AH+, and AH- compared to controls. DUDIT scores were higher in patients, AH+, and AH- compared to controls. GAF-F and GAF-S scores were lower in AH+ compared to AH-. Positive, excited, and depressed symptom factors, and CPZ-equivalent doses were higher in AH+ compared to AH-. See **Table 1** for clinical and demographic variables for the final sample and **Supplementary Tables 1** and **2** for the same divided by dataset.

### 3.2 Tract- and metric-specific analyses

See **Figure 2** for bar plots of Cohen’s d effect sizes for MD and RD, **Supplementary Figure 7** for bar plots of Cohen’s d effect sizes for FA and AD, and **Supplementary Table 4-7** for Cohen’s d effect sizes and FDR-adjusted and unadjusted p-values for each DTI metric.

**Figure 2.**
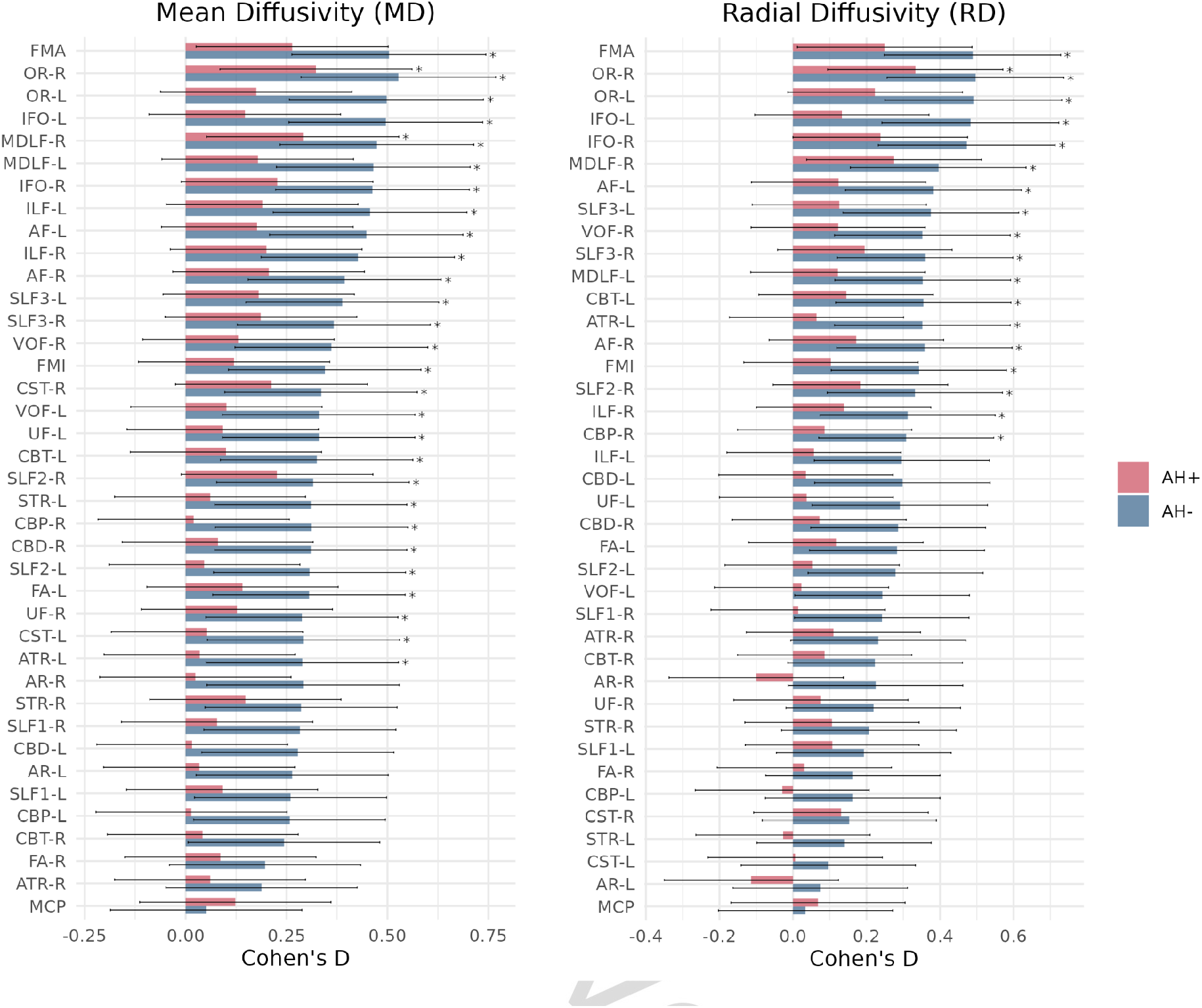
Cohen’s d effect sizes for the contrast between patients with schizophrenia with (AH+) and without (AH-) current hallucinations and healthy controls for radial diffusivity (RD) and mean diffusivity (MD). Fibre tracts are ordered by estimated effect size. Error bars indicate 95% confidence intervals. Significant differences after FDR correction are marked with asterisks.

We observed no significant group differences between AH+ and AH- and healthy controls for FA or AD. Compared to controls, AH- had higher MD in several fibre tracts, including the bilateral AF, right AR, left ATR, right CBD, right CBP, left CBT, bilateral CST, left FA, bilateral ILF, bilateral IFO, bilateral MDLF, bilateral OR, bilateral STR, right SLF1, bilateral SLF2, bilateral SLF3, bilateral UF, bilateral VOF, FMA, and FMI. We also observed higher MD in AH+ in the right MDLF and the right OR compared to controls.

Higher RD was observed in AH- for the bilateral AF, left ATR, right CBP, left CBT, right ILF, bilateral IFO, bilateral MDLF, bilateral OR, right SLF2, bilateral SLF3, right VOF, FMA, and FMI compared to controls. We also observed higher RD in the right OR in AH+ compared to controls. There were no significant differences between AH+ and AH- after correction for multiple testing for any of the DTI metrics.

We found widespread group differences between patients with and without a lifetime history of AH and controls for MD and RD, but not AD and FA. See **Supplementary Note 4** for details on these results. When comparing the whole patient group (SCZ) with controls, significant differences were found in a wide range of fibre tracts for MD, RD, and AD, but not FA. See **Supplementary Figure 8** and **9** for bar plots of Cohen’s d effect sizes, **Supplementary Tables 8-11**, and **Supplementary Note 5** for details on these results.

### 3.3 Uni- and multimodal general factor analyses

Kaiser-Meyer-Olkin test and Bartlett’s test of sphericity indicated excellent suitability for factor analysis. See **Supplementary Figures 10-13** for correlation matrices between fibre tracts and **Supplementary Figure 14** for a density plot of the correlations.

Explained variance of the unimodal g-factors ranged from 41.1% for g-FA to 63.9% for g-MD, whereas the second principal components only explained between 5.4% for g-RD to 6.6% for g-AD. See **Supplementary Figure 15** for scree and variable contribution plots and **Supplementary Table 12** for tract-wise correlations with each general factor.

For the multimodal g-factors, the explained variance was 64.3% for g-Dim1 and 32.5% for g-Dim2. Correlations with g-Dim1 were negative (r = -0.72) for FA, and positive for MD (r = 0.95), RD (r = 0.99), and AD (r = 0.42). For g-Dim2 the correlations were negative for FA (r = -0.66), MD (r = -0.25), and AD (r = 0.88), and positive for RD (r = 0.13). Most of the variance of g-Dim1 was contributed by RD (37.8%), MD (35.3%), and FA (19.9%). For g-Dim2, AD (60.1%) and FA (33.9%) contributed the most to the variance. See **Supplementary Table 13** for correlations and contributions of each DTI metric to the multimodal g-factors and **Supplementary Figure 16** for scree plots and variable contribution plots.

In the regression analyses we found significant group differences between AH- and controls for all uni- and multimodal g-factors, except g-FA. These associations indicated lower g-Dim2 and higher g-MD, g-RD, and g-AD in AH- compared to controls. No significant differences between AH+ and controls were observed for any of the g-factors. See **Figure 3** for violin plots of each g-factor for each group adjusted for age, age^2^, and sex.

**Figure 3.**
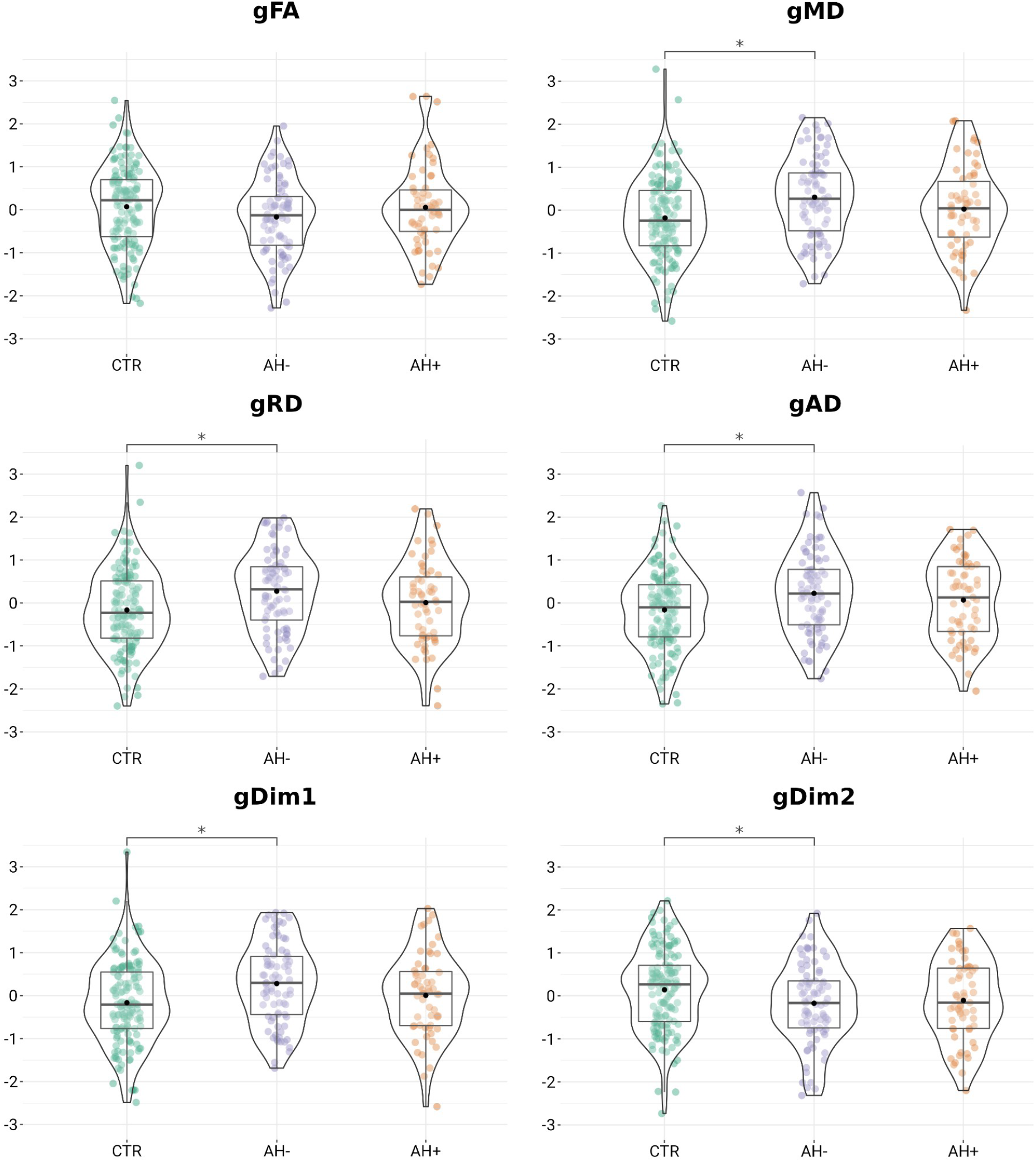
Uni- and multimodal g-factors residualised with respect to age, and age^2^, and sex for controls (CTR), patients with current hallucinations (AH+) and patients without current hallucinations (AH-).

### 3.4 Associations with clinical and demographic variables

We found no significant associations between uni- and multimodal g-factors and age at onset, duration of illness, antipsychotic medication use (yes/no), CPZ-equivalent dose, or the positive, negative, disorganised, depressed, or excited Wallwork symptom factors.

ICV was significantly associated with g-FA, g-RD, g-AD, and g-Dim2. Group differences between AH- and healthy controls remained significant for g-RD, g-AD, g-Dim2 when also adjusting for ICV. There were no significant associations between g-factors and BMI or significant interactions between current AH status and age or sex for any of the g-factors.

## 4 Discussion

The main finding of the present study was a widespread pattern of tract-wise differences represented by higher MD and RD in patients with SCZ *without* current AH (AH-) compared to healthy controls. Surprisingly, only MD in the right middle longitudinal fasciculus (MDLF) and both MD and RD in the right optic radiation (OR) differed significantly in patients with current AH (AH+) relative to healthy controls, indicating higher MD and RD. In line with these findings, uni- and multimodal g-factors differed only between the AH- group and healthy controls. These findings suggest that altered DTI metrics in schizophrenia are not a specific feature underlying AH and instead point toward a more complex relationship between WM microstructure and AH.

Most previous DTI studies on AH in SCZ have focused on a limited set of fibre tracts under the assumption that WM alterations relevant to AH are localised within the language and auditory processing circuitry (LAPC). We observed higher MD and RD in several fibre tracts within the LAPC in patients without current AH compared to controls. These fibre tracts included the arcuate fasciculus (AF), inferior longitudinal fasciculus (ILF), the superior longitudinal fasciculus (SLF), inferior fronto-occipital fasciculus (IFO), and the uncinate fasciculus (UF). We also found associations with AH in interhemispheric fibre tracts including the forceps major and minor (FMA and FMI) and the dorsal, peri-genual and temporal subsections of the cingulum bundle (CBD, CBP, and CBT), as well as in fibre tracts not in the LAPC e.g., the vertical occipital fasciculus (VOF) and the OR. The involvement of interhemispheric pathways in AH in SCZ has been proposed previously, but findings have been inconsistent (Curcic-Blake et al., 2015; Knöchel et al., 2012; Leroux, Delcroix, & Dollfus, 2017; Shao et al., 2021; Wigand et al., 2015). Our results provide evidence for the involvement of interhemispheric fiber tracts in AH and suggest that the relationship between WM microstructure and AH is widespread rather than confined to the LAPC.

The AH- group differed significantly compared to healthy controls for all uni- and multimodal g-factors, except g-FA, suggesting a generalised effect. In agreement with the literature (Chamberland et al., 2019; Geeraert et al., 2020; Vaher et al., 2022), the first principal components explained most of the variation in the unimodal principal component analyses (41.1% to 63.9%), while the second principal components explained only 5.4% to 6.6%. For the multimodal g-factors, the second principal component explained a non-negligible proportion of the variation, which is also in line with past studies (Chamberland et al., 2019; Geeraert et al., 2020; Vaher et al., 2022). Interestingly, g-AD was significantly higher in AH- patients compared to controls although there were no significant tract-specific differences for AD. Similarly, g-Dim2, which mostly received contributions from FA (33.9%) and AD (60.1%), was significantly lower in AH- compared to controls. These findings may indicate enhanced sensitivity to WM microstructure differences with the use of dimensionality reduction.

In a study on individuals at clinical high risk for psychosis and patients with first-episode psychosis, Sato et al. (2021) found positive associations between hallucination severity and MD in the left SLF and inferior IFO. Though this contrasts with our findings, the patients included in our study had a relatively long duration of illness (mean=10.6 years). Given the dynamic nature of WM microstructure (Beck et al., 2021), it would be necessary to perform longitudinal data collection to assess trajectories of WM microstructure and AH. We could not ascertain if the observed group differences emerged after illness onset or point towards a subgroup of patients that are less prone to AH and more likely to exhibit WM differences as measured with DTI. Notably, our analyses on lifetime AH showed higher MD and RD in patients with lifetime AH compared to controls including the left AF and left ILF and the bilateral IFO. We encourage future studies to assess the longitudinal course of WM microstructure and relationships with AH as the illness progresses from an acute to a more chronic phase.

There were no significant group differences in g-factors between patients who used antipsychotic medication compared to patients who did not, nor significant associations between g-factors and CPZ-equivalent dose. However, only a small number of patients did not use antipsychotic medication and we did not have data on cumulative exposure which may have challenged our ability to detect medication-related effects. The literature on the effects of antipsychotic medication on DTI metrics is sparse and has been largely focused on FA (Sagarwala & Nasrallah, 2021). Previous studies have reported both reduced (Wang et al., 2013) and increased (Ozcelik-Eroglu et al., 2014; Serpa et al., 2017; Zeng et al., 2016) FA following antipsychotic treatment. Further studies with more comprehensive medication data are needed to characterise the effects of antipsychotic medication on WM microstructure.

Strengths of the study included a clinically well-characterised patient group, which allowed us to test associations with current antipsychotic medication use and dose, duration of illness, and psychotic symptoms. Importantly, our relatively large sample size afforded us the statistical power to assess a broader range of fibre tracts than previous studies and to combine and compare FA, MD, RD, and AD through dimensionality reduction. Fibre tractography was performed using a reproducible framework at the subject-level rather than relying on maps from subject space to a common template. This improves the accuracy of fibre tract reconstruction by taking individual anatomical differences into account.

Some limitations also apply. Since the data were cross-sectional, we could not estimate longitudinal changes to WM microstructure. We also did not have access to cumulative exposure to antipsychotic medication. The current AH+ group was smaller than the current AH- group which may have reduced sensitivity. In line with some previous studies (Catani et al., 2011; Leroux et al., 2017; Xi et al., 2016; Xie et al., 2019), we only observed significant differences compared to controls and not between AH+ and AH-. As such, strong conclusions on the differences between AH+ and AH- should be avoided. Furthermore, while we matched patients and controls on scanner and corrected for scanner effects using a well-established harmonisation procedure, we cannot rule out that residual effects of scanner remained. Finally, DTI metrics are indirect measures of WM microstructure and strong biophysical conclusions should be avoided (Figley et al., 2022).

In conclusion, we found higher MD and RD across widespread fibre tracts mainly in patients without current AH compared to controls. In contrast to our hypothesis, patients with SCZ and current AH only differed significantly relative to healthy controls for MD in the MDLF and the OR and RD in the OR. These results challenge the idea that altered DTI metrics in the LAPC in patients with SCZ is a specific feature underlying AH. Instead, the findings suggest a more complex relationship between AH status and WM microstructure. We encourage future studies to investigate the longitudinal course of AH and WM microstructure and to employ more direct measures of myelin alongside detailed clinical evaluations of AH.

## Data Availability

Given a formal data sharing agreement and approval from the local ethics committees, access to the data presented in this study can be provided upon reasonable request to the authors.

## Acknowledgements

We are grateful for the study participants who were willing to participate in this project and the clinicians who were involved in recruitment and assessments at NORMENT and as part of the HUBIN project. Data services were provided by the Services for Sensitive Data (TSD) facilities, operated and developed by the TSD service group in the University of Oslo IT services department (USIT).

## Ethics declaration

The studies were carried out in accordance with the Helsinki Declaration and all participants provided written informed consent. The TOP project was approved by the Regional Committee for Medical Research Ethics and the Norwegian Daxsta Inspectorate. The HUBIN project was approved by the Swedish Ethical Review Authority at Karolinska Institutet. Data handling complied with Norwegian Data Protection Authority and GDPR regulations.

