## Supplementary Materials for "Widespread alterations of diffusion tensor imaging metrics in patients with schizophrenia without current auditory hallucinations"

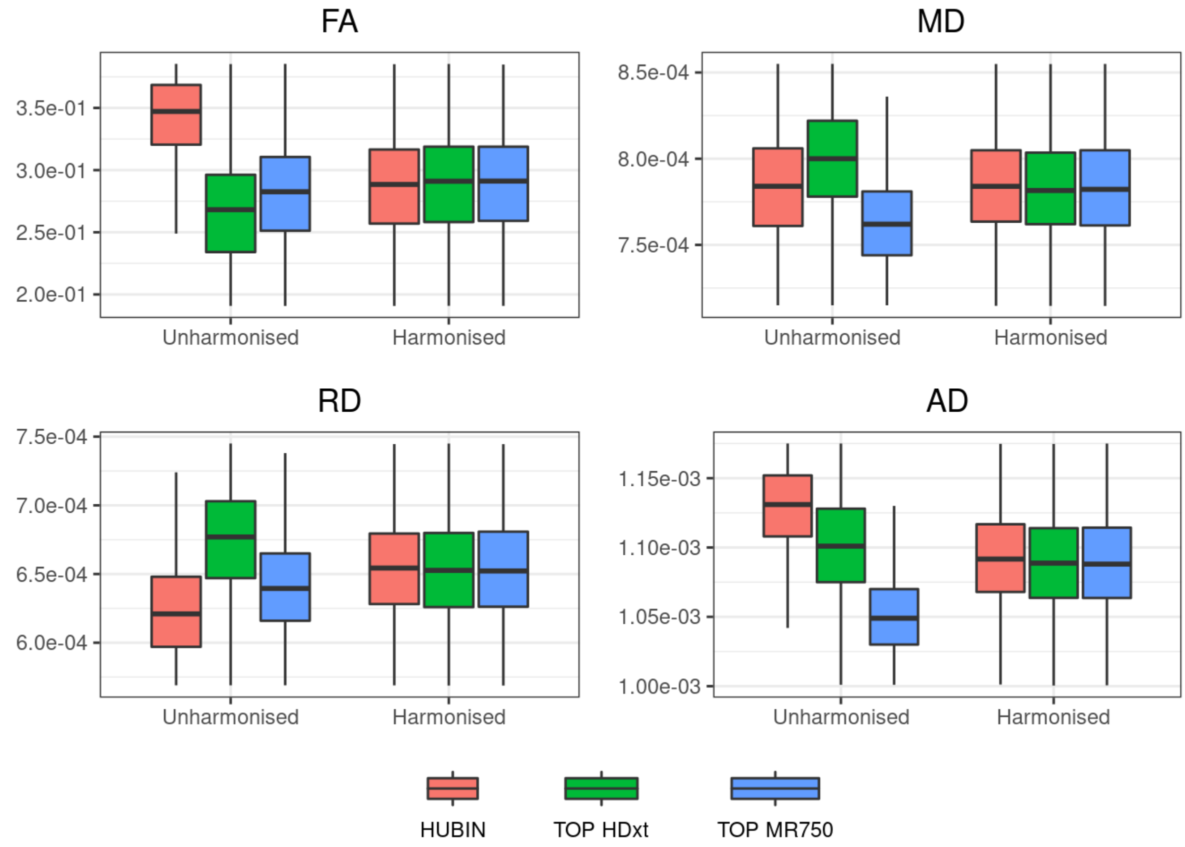


**Supplementary Figure 1.** Boxplots of mean DTI measurements across all 39 fibre tracts before and after scanner harmonisation.

ersion

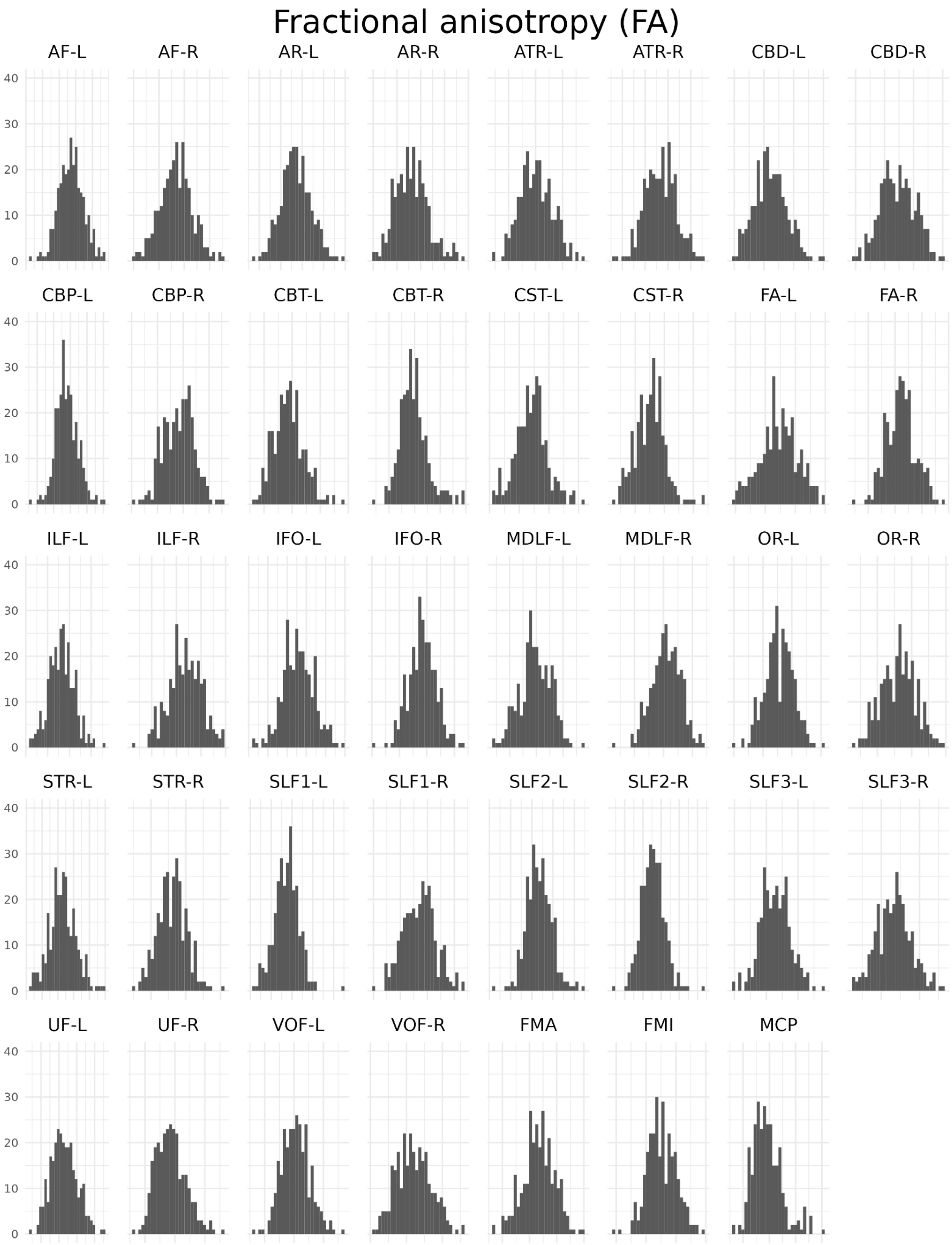


**Supplementary Figure 2.** Histograms for each fibre tract for fractional anisotropy (FA) after scanner harmonisation.

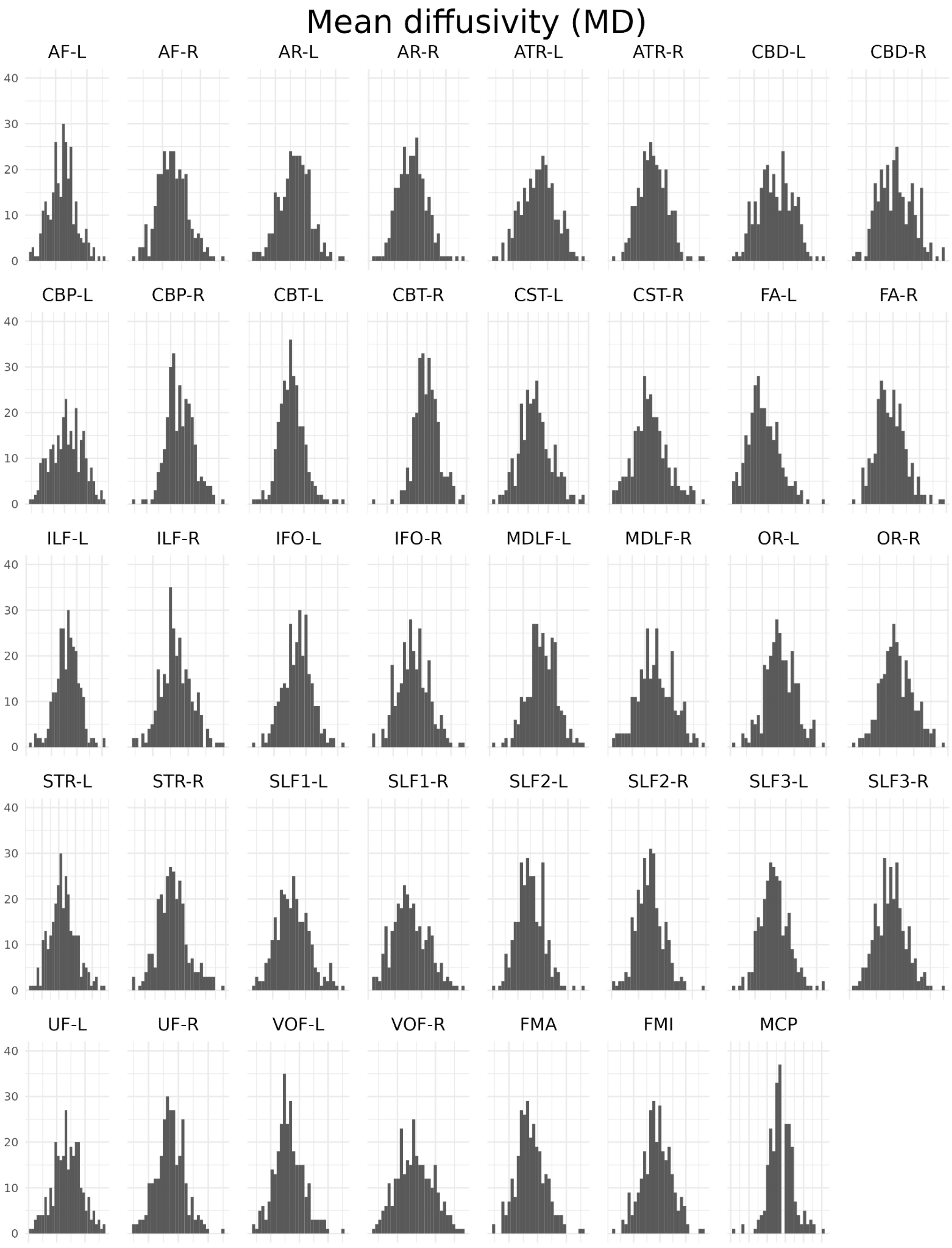


**Supplementary Figure 3.** Histograms for each fibre tract for mean diffusivity (MD) after scanner harmonisation.

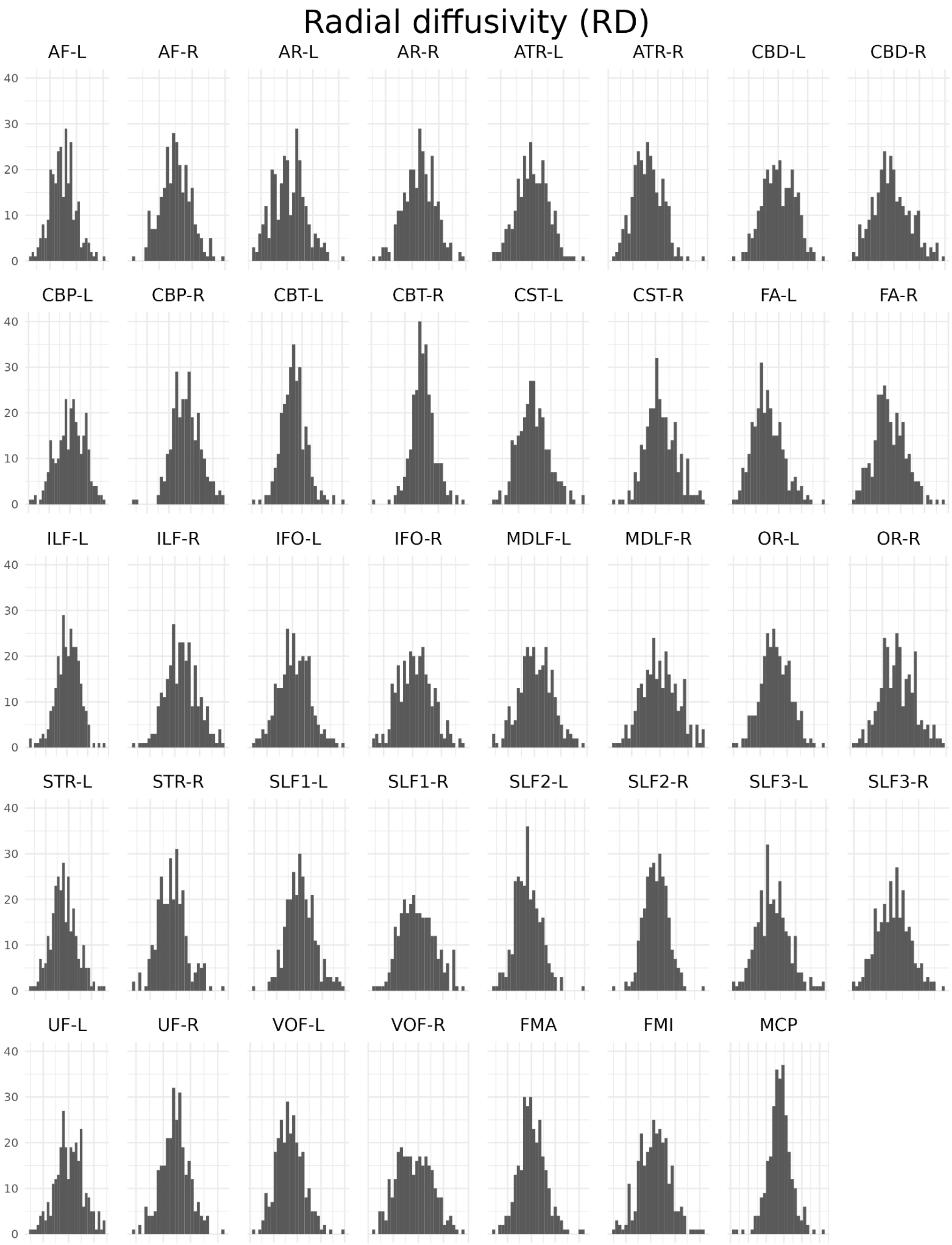


**Supplementary Figure 4.** Histograms for each fibre tract for radial diffusivity (RD) after scanner harmonisation.

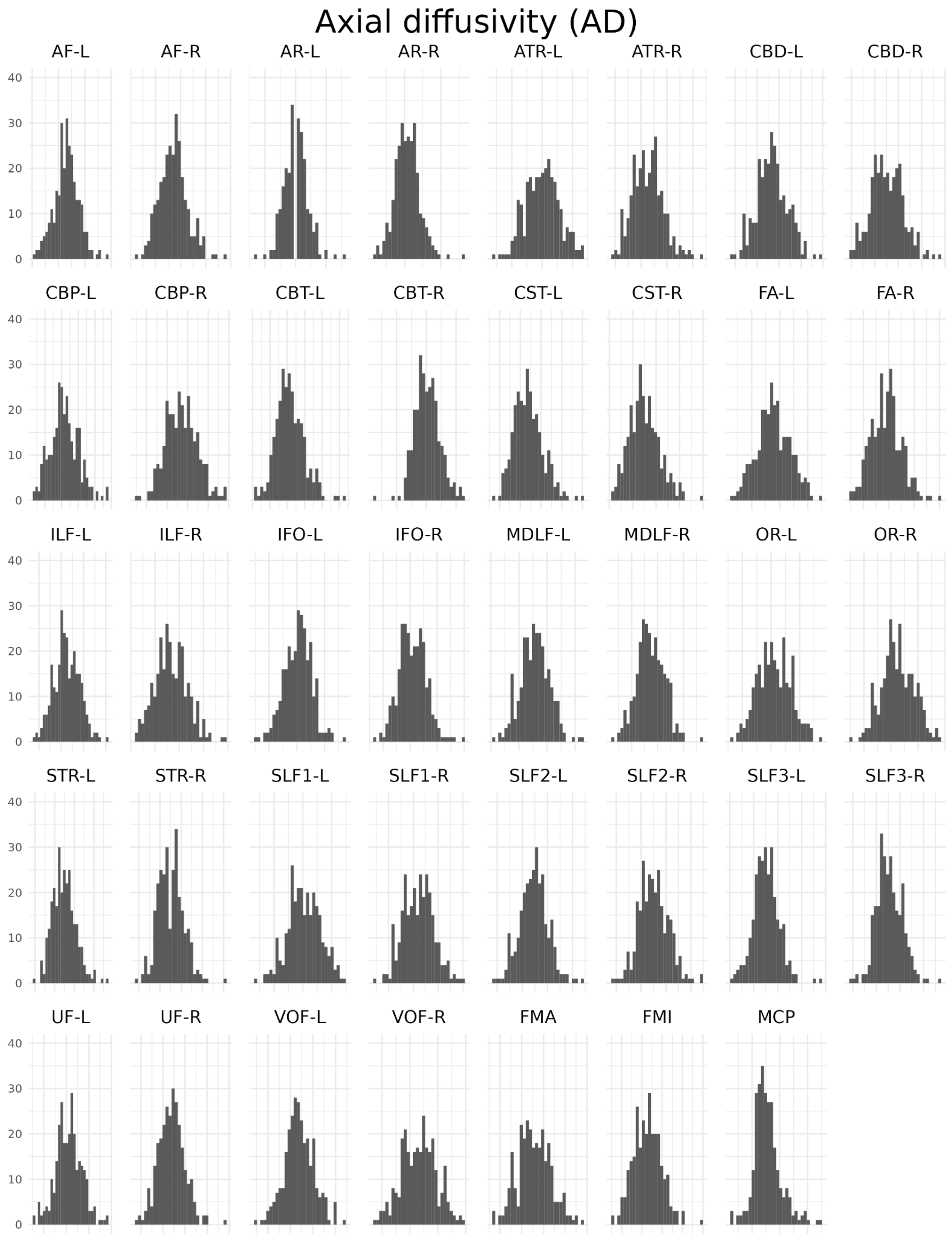


**Supplementary Figure 5.** Histograms for each fibre tract for axial diffusivity (AD) after scanner harmonisation.

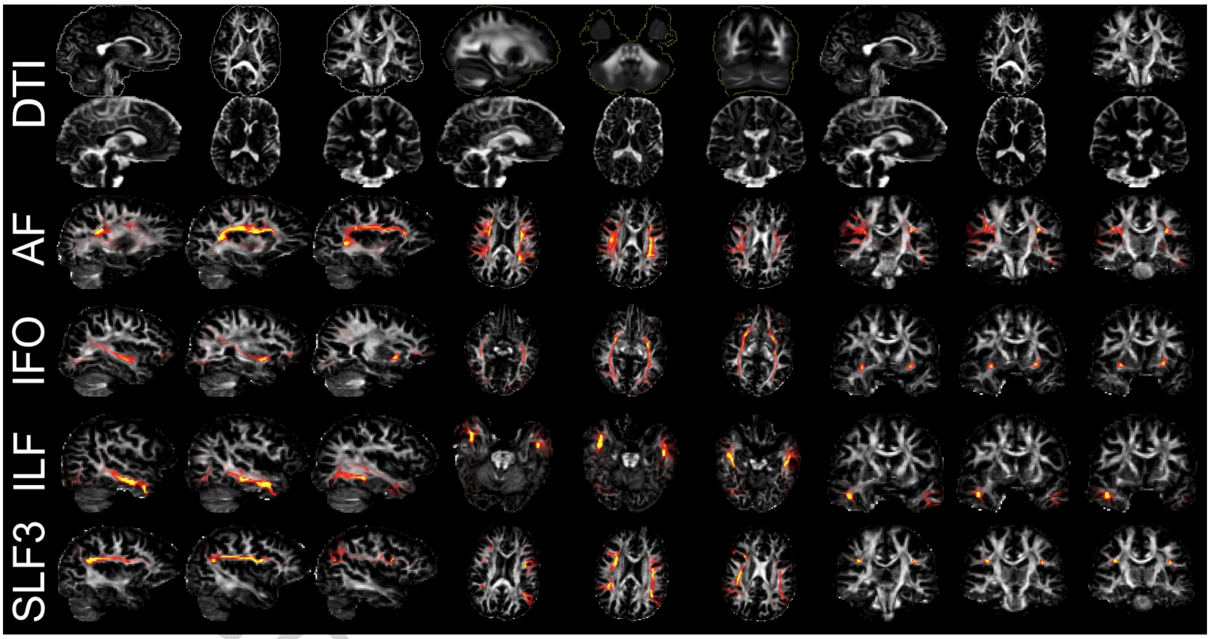


**Supplementary Figure 6.** A selection of reconstructed fibre tracts for a randomly chosen participant. The first row shows fractional anisotropy (FA) maps with whole-brain segmentation from BET (white outline), whole-brain segmentation mapped from the standard MNI152 template (yellow outline), and without whole-brain masks. The second row shows radial diffusivity (RD), axial diffusivity (AD), and mean diffusivity (MD). The three bottom rows show probabilistic segmentations of the arcuate fasciculus (AF), the inferior fronto-occipital fasciculus (IFO), the inferior longitudinal fasciculus (ILF), and the third branch of the superior longitudinal fasciculus (SLF3).

print version

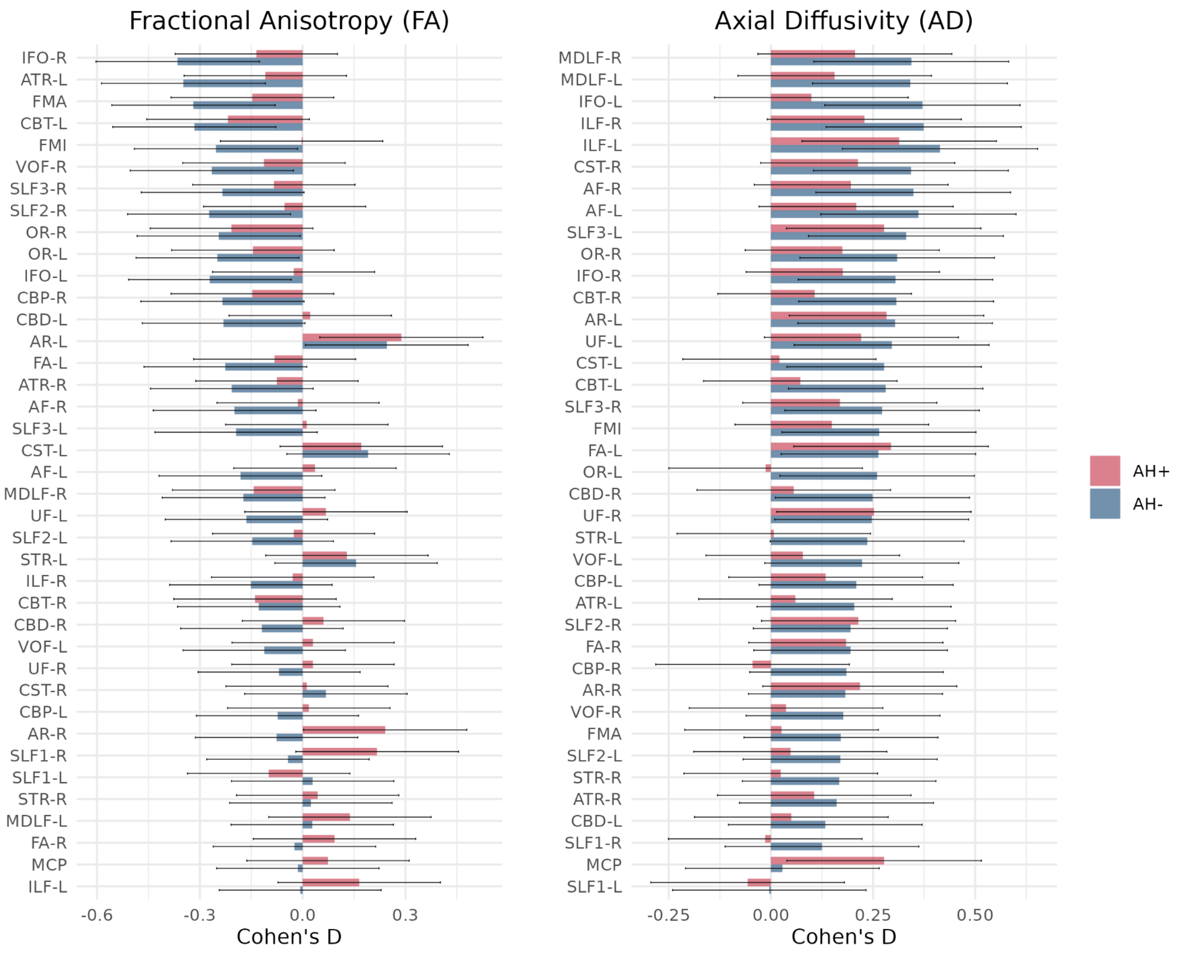


**Supplementary Figure 7.** Cohen's d effect sizes for the contrast between patients with schizophrenia with (AH+) and without (AH-) current hallucinations and healthy controls for fractional anisotropy (FA) and axial diffusivity (AD). Fibre tracts are ordered by effect size. Error bars indicate 95% confidence intervals.

version

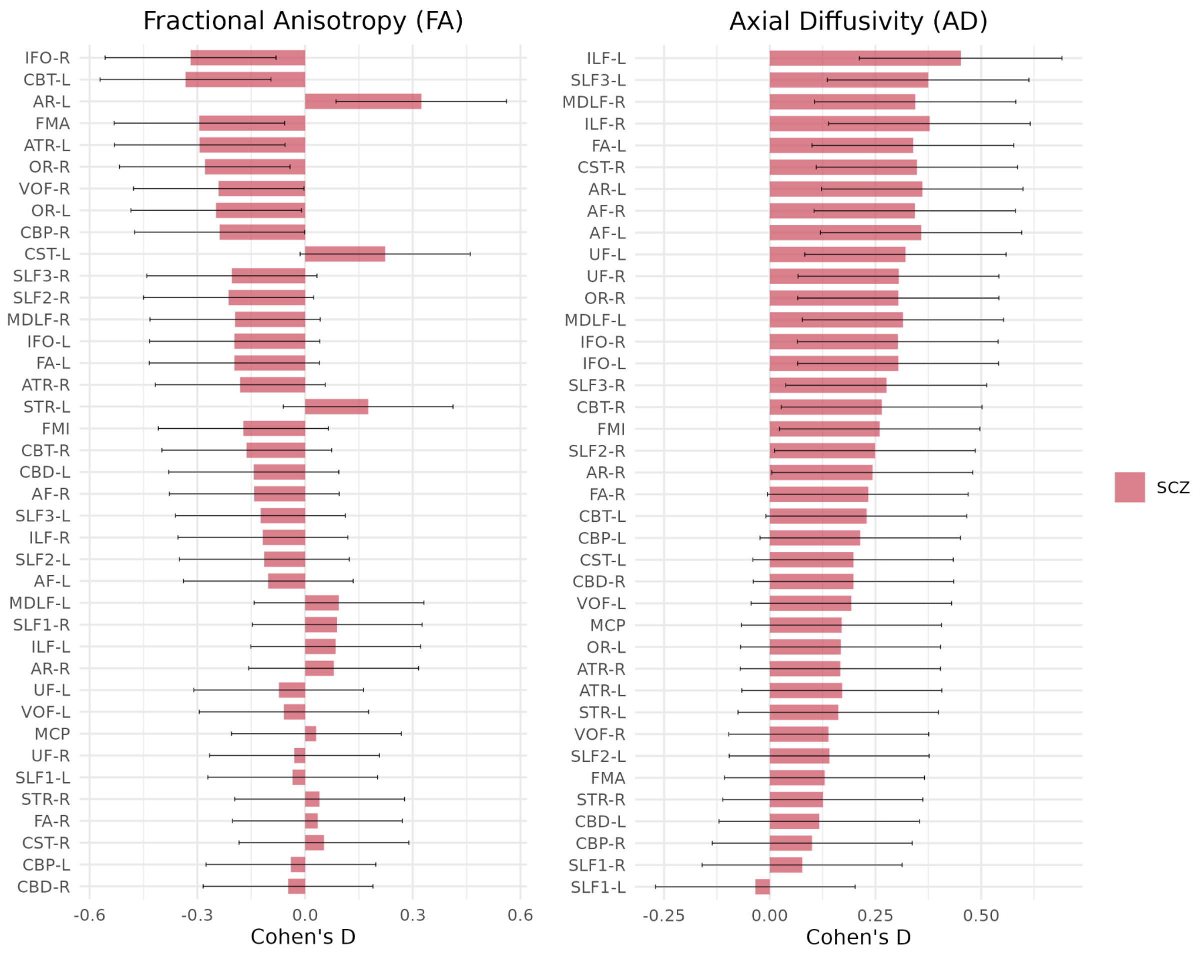


version

**Supplementary Figure 8.** Cohen's d effect sizes for the contrast between patients with schizophrenia (SCZ) and healthy controls for fractional anisotropy (FA) and axial diffusivity (AD). Fibre tracts are ordered by effect size. Error bars indicate 95% confidence intervals.

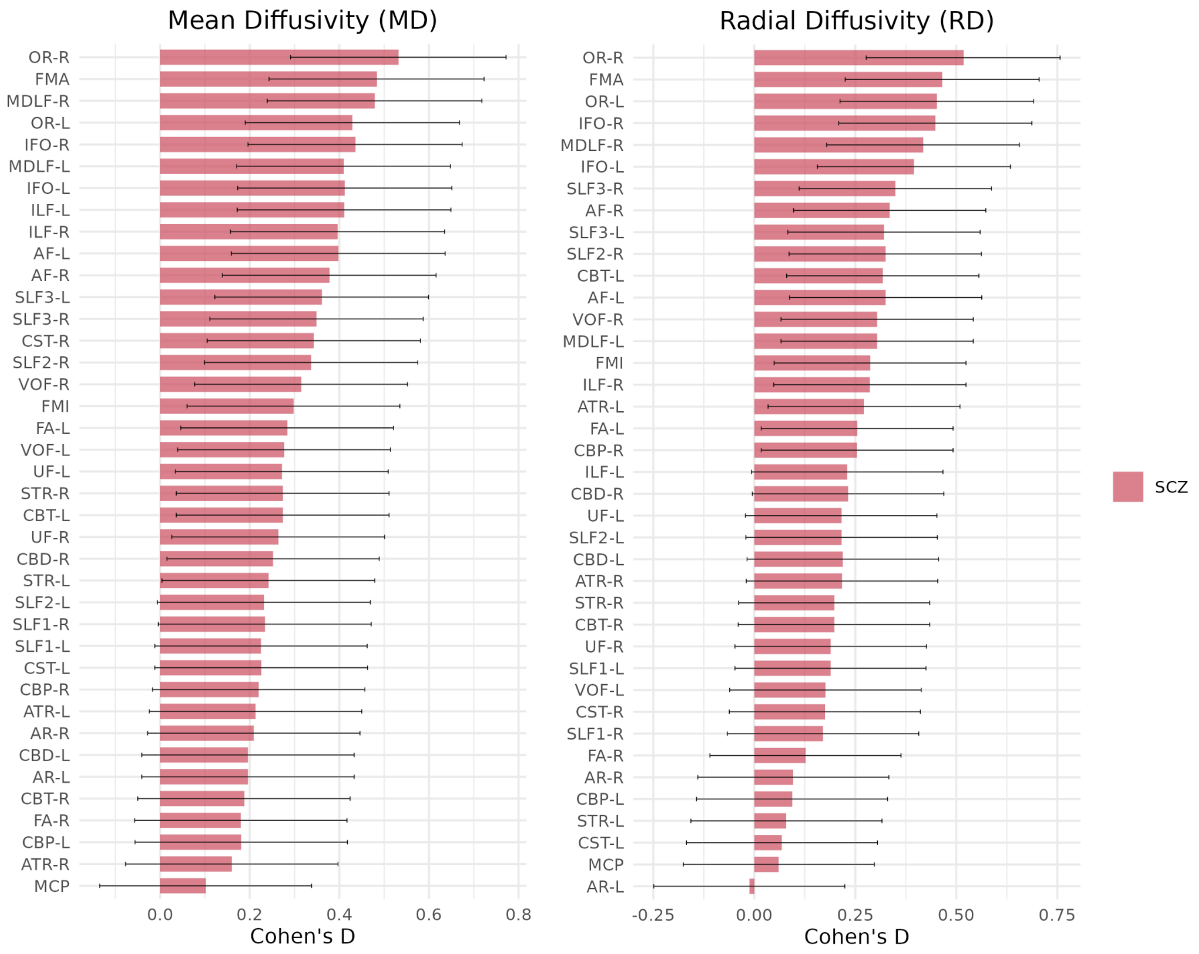


ersion

**Supplementary Figure 9.** Cohen's d effect sizes for the contrast between patients with schizophrenia (SCZ) and healthy controls for mean diffusivity (MD) and radial diffusivity (RD). Fibre tracts are ordered by effect size. Error bars indicate 95% confidence intervals.

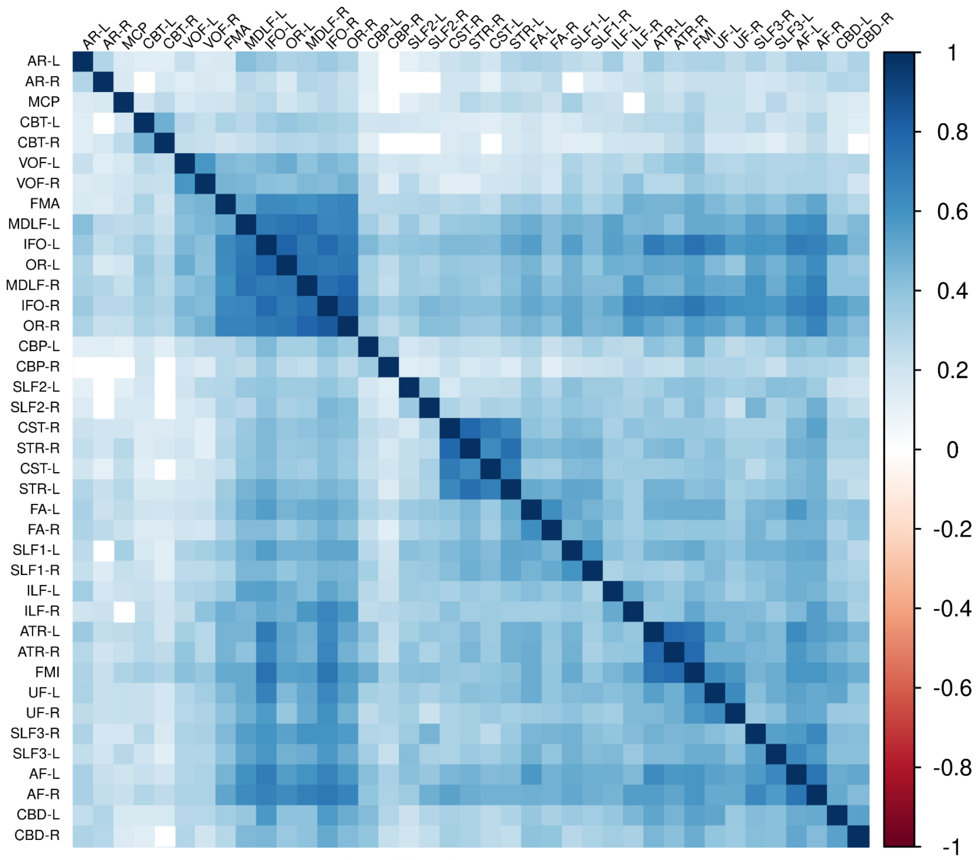


**Supplementary Figure 10.** Pearson correlations between each the measurement in fibre tract for fractional anisotropy (FA) sorted according to the hierarchical cluster order.

version

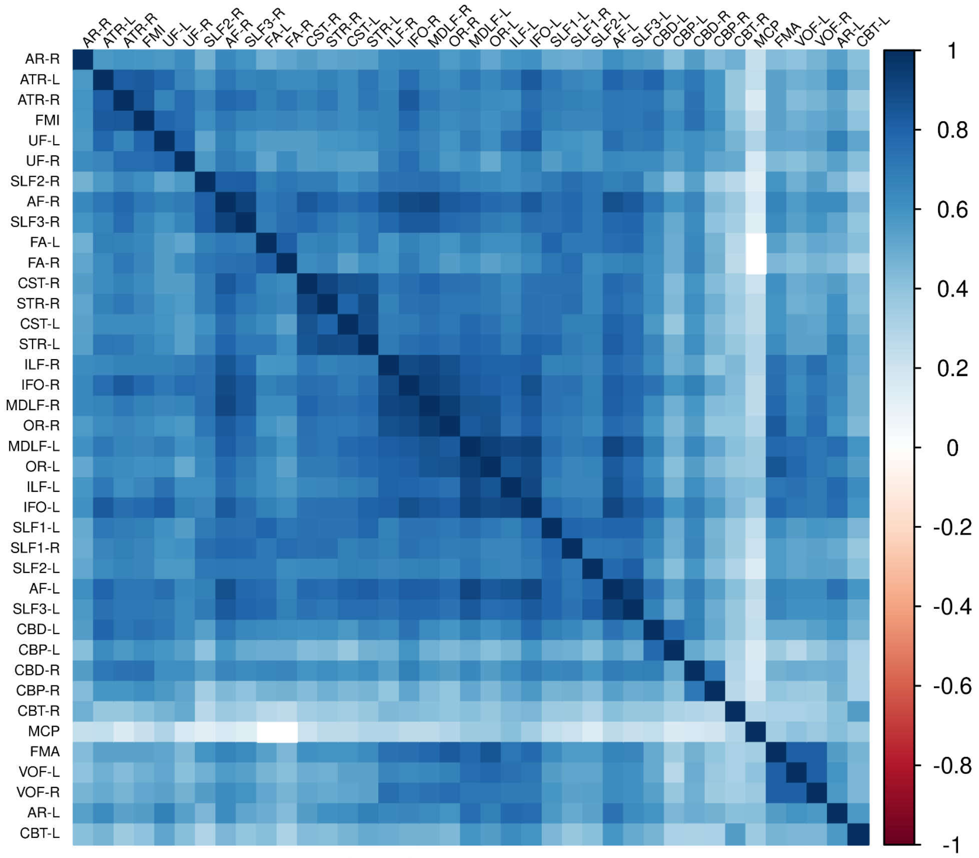


**Supplementary Figure 11.** Pearson correlations between each the measurement in fibre tract for mean diffusivity (MD) sorted according to the hierarchical cluster order.

version

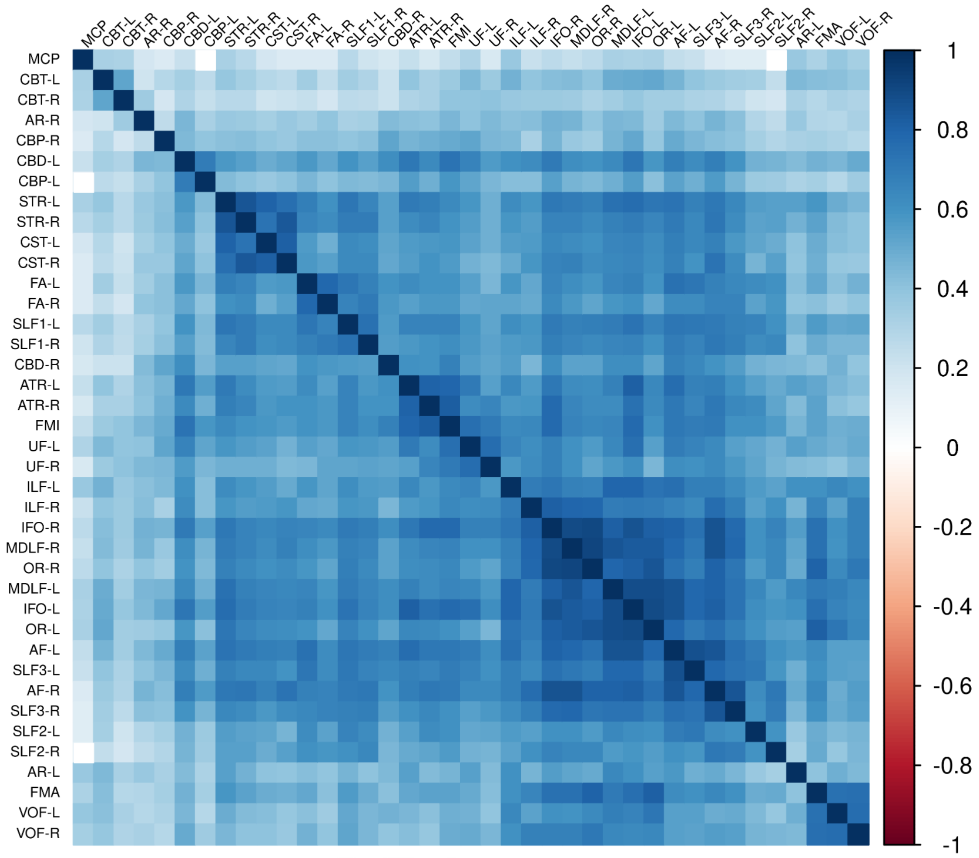


**Supplementary Figure 12.** Pearson correlations between each the measurement in fibre tract for radial diffusivity (RD) sorted according to the hierarchical cluster order.

version

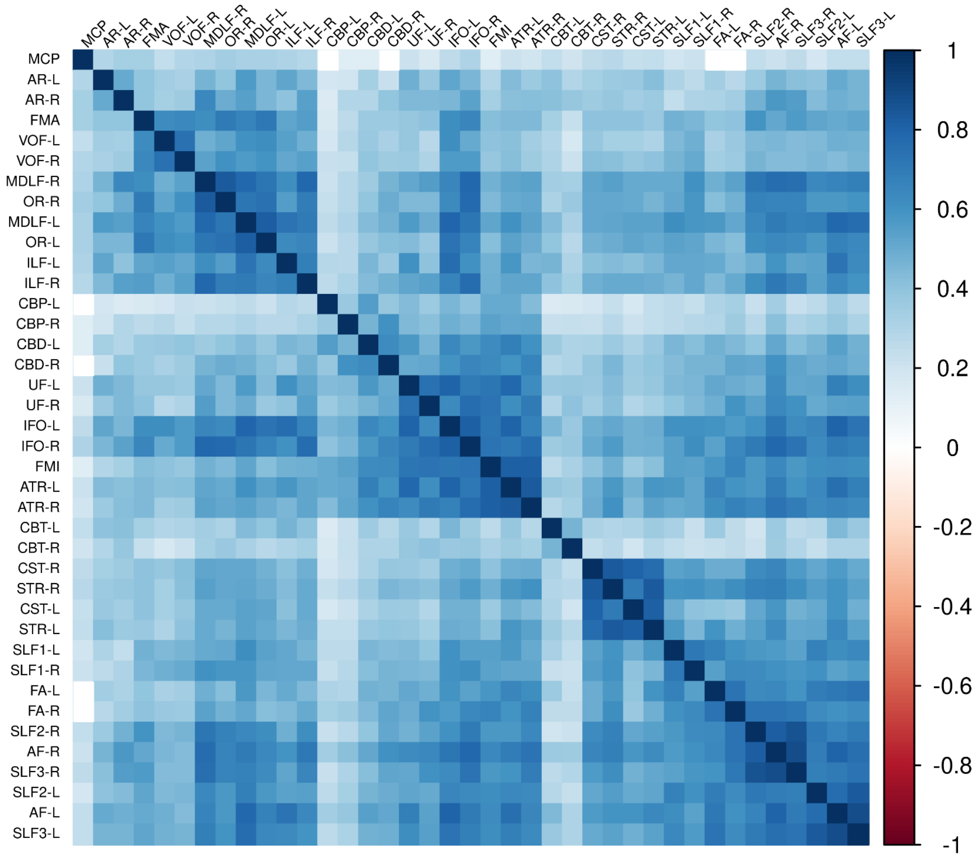


**Supplementary Figure 13.** Pearson correlations between each the measurement in fibre tract for axial diffusivity (AD) sorted according to the hierarchical cluster order.

version

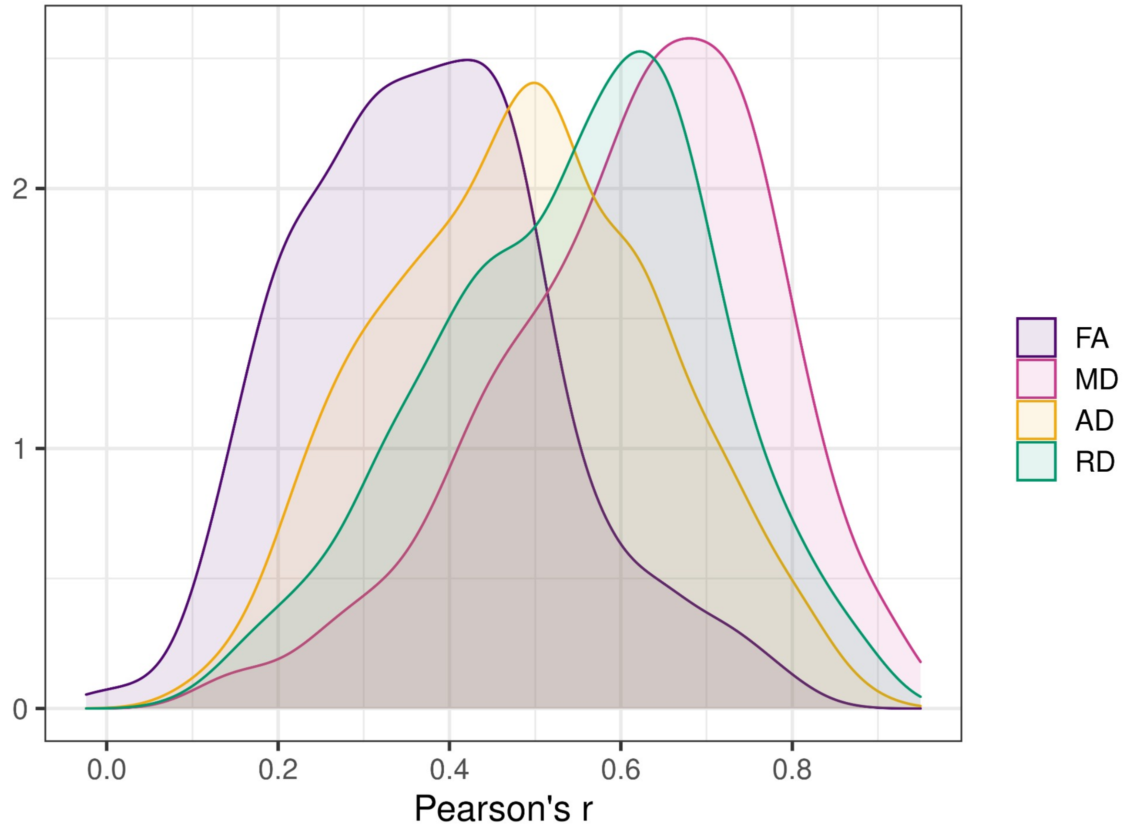


version

**Supplementary Figure 14.** Distribution of Pearson correlations between fibre tracts for all DTI metrics.

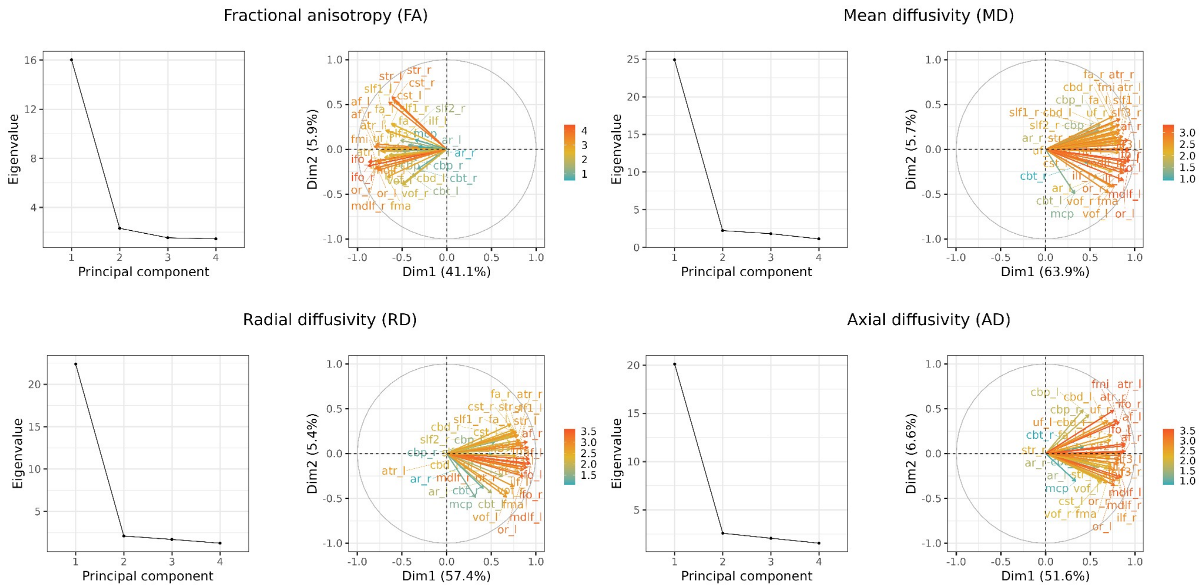


**Supplementary Figure 15.** Scree plots and variable contribution plots for each of the unimodal principal component analyses.

print version

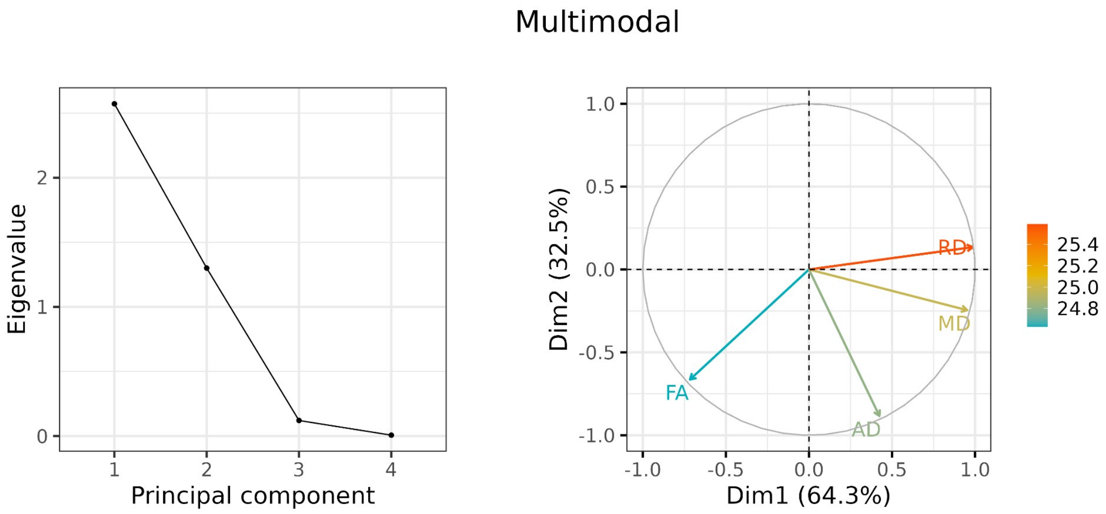


**Supplementary Figure 16.** Scree plot and variable contribution plot for the multimodal principal component analysis.

Preprint version

| | CTR (N=114) | SCZ (N=114) | AH+ (N=44) | AH- (N=70) |
|---|---|---|---|---|
| **Demographics and IQ** | | | | |
| **Age [years]** | 29.0 (7.9) | 29.1 (8.3) | 28.9 (8.6) | 29.3 (8.2 |
| **Sex [female]** | 44 (38.6%) | 44 (38.6%) | 17 (38.6%) | 27 (38.6%) |
| **Education [years]** | 14.1 (1.9) | 12.3 (2.2) | 12.0 (2.5) | 12.5 (2.0) |
| **Handedness [right]** | 46 (85.2%) | 50 (96.2%) | 21 (95.5%) | 29 (96.7%) |
| **BMI** | 24.6 (3.6) | 25.9 (5.3) | 25.2 (4.9) | 26.4 (5.6) |
| **IQ** | 113.4 (10.4) | 101.8 (13.3) | 101.8 (13.6) | 101.8 (13.2) |
| **Alcohol and drug use** | | | | |
| **AUDIT** | 5.7 (3.2) | 6.7 (6.3) | 7.7 (7.5) | 6.1 (5.3) |
| **DUDIT** | 0.3 (1.3) | 5.1 (8.1) | 6.4 (9.1) | 4.3 (7.3) |
| **Clinical variables** | | | | |
| **Age at onset [years]** | N.A. | 22.0 (6.6) | 20.7 (5.5) | 22.8 (7.2) |
| **Duration of illness [years]** | N.A. | 7.0 (6.9) | 7.8 (8.0) | 6.5 (6.3) |
| **Psychiatric hospital admissions** | N.A. | 2.9 (3.3) | 3.1 (3.3) | 2.8 (3.3) |
| **GAF-F** | N.A. | 47.8 (13.0) | 41.9 (10.6) | 51.5 (13.1) |
| **GAF-S** | N.A. | 48.6 (13.3) | 41.8 (9.2) | 52.9 (13.8) |
| **PANSS factors** | | | | |
| **Positive** | N.A. | 9.0 (4.0) | 12.4 (2.5) | 6.8 (3.1) |
| **Negative** | N.A. | 13.2 (5.1) | 14.0 (5.6) | 12.7 (4.8) |
| **Disorganised** | N.A. | 5.6 (2.9) | 6.2 (3.5) | 5.3 (2.3) |
| **Excited** | N.A. | 5.3 (2.1) | 6.0 (2.9) | 4.9 (1.1) |
| **Depressed** | N.A. | 7.8 (2.9) | 8.7 (2.6) | 7.3 (2.9) |
| **Medication use** | | | | |
| **AP use [yes]** | N.A. | 104 (92.0%) | 38 (88.4%) | 66 (94.3%) |
| **CPZ-equiv. AP dose [mg/day]** | N.A. | 328.8 (196.8) | 419.1 (248.4) | 275.2 (134.2) |
| **Antiepileptic use* [yes]** | N.A. | 14 (12.6%) | 4 (9.5%) | 10 (14.5%) |
| **Antidepressant use* [yes]** | N.A. | 29 (26.1%) | 13 (31.0%) | 16 (23.2%) |

**Supplementary Table 1.** Demographic and clinical characteristics of the healthy controls, the patient group of the TOP dataset and the patient subgroups with (AH+) and without (AH-) current hallucinations. Continuous variables are reported as mean (standard deviation) and categorical variables are given as count (proportion).

| | CTR (N=26) | SCZ (N=26) | AH+ (N=15) | AH- (N=11) |
|---|---|---|---|---|
| **Demographics and IQ** | | | | |
| **Age [years]** | 51.6 (8.0) | 51.5 (8.0) | 49.0 (7.6) | 54.7 (7.7) |
| **Sex [female]** | 6 (23.1%) | 6 (23.1%) | 3 (20.0%) | 3 (27.3%) |
| **Education [years]** | 14.4 (3.8) | 12.3 (2.6) | 12.9 (2.8) | 11.5 (2.3) |
| **Handedness [right]** | 23 (92.0%) | 20 (80.0%) | 12 (80.0%) | 8 (80.0%) |
| **Clinical variables** | | | | |
| **Age at onset [years]** | N.A. | 23.0 (4.7) | 22.2 (3.8) | 24.5 (6.0) |
| **Duration of illness [years]** | N.A. | 27.7 (7.5) | 26.8 (7.0) | 29.4 (8.8) |
| **GAF-F** | N.A. | 45.2 (6.1) | 46.0 (6.9) | 44.1 (4.9) |
| **GAF-S** | N.A. | 43.1 (5.8) | 41.7 (4.5) | 45.0 (7.1) |
| **Medication use** | | | | |
| **AP use [yes]** | N.A. | 25 (96.2%) | 14 (93.3%) | 11 (100.0%) |
| **CPZ-equiv. AP dose [mg/day]** | N.A. | 421.9 (230.5) | 453.9 (284.9) | 378.3 (124.5) |

**Supplementary Table 2.** Demographic and clinical characteristics of the healthy controls, the patient group of the HUBIN dataset and the patient subgroups with (AH+) and without (AH-) current hallucinations. Continuous variables are reported as mean (standard deviation) and categorical variables are given as count (proportion).

Preprint version

| | OUS, Norway | OUS, Norway | KI, Sweden |
|---|---|---|---|
| **Scanner model** | GE Signa HDxt | GE Discovery MR750 | GE Discovery MR750 |
| **Field strength (Tesla)** | 3T | 3T | 3T |
| **Repetition time (ms)** | 15000 | 8150 | 6000 |
| **Echo time (ms)** | 85 | 83 | 83 |
| **Flip angle (°)** | 90 | 90 | 90 |
| **Resolution ($mm^3$)** | 1.875 × 1.875 × 2.5 | 2 × 2 × 2 | 0.94 × 0.94 × 2.9 |
| **Diffusion directions** | 30 | 60 | 60 |
| **b-value ($s/mm^2$)** | 1000 | 1000 | 1000 |
| **Number of b=0 images** | 2 | 5 | 10 |

**Supplementary Table 3.** Diffusion imaging acquisition parameters for each dataset.

Preprint version

| | Current AH- | | | | | Current AH+ | | |
|---|---|---|---|---|---|---|---|---|
| Fibre tract | Cohen's d | p-value | p-value (FDR) | | Fibre tract | Cohen's d | p-value | p-value (FDR) |
| **AF-L** | -0.18 | 0.134 | 0.419 | | **AF-L** | 0.04 | 0.767 | 0.937 |
| **AF-R** | -0.20 | 0.101 | 0.357 | | **AF-R** | -0.01 | 0.911 | 0.946 |
| **AR-L** | 0.25 | 0.043 | 0.279 | | **AR-L** | 0.29 | 0.018 | 0.277 |
| **AR-R** | -0.08 | 0.531 | 0.798 | | **AR-R** | 0.24 | 0.047 | 0.279 |
| **ATR-L** | -0.35 | 0.004 | 0.166 | | **ATR-L** | -0.11 | 0.368 | 0.638 |
| **ATR-R** | -0.21 | 0.088 | 0.329 | | **ATR-R** | -0.08 | 0.534 | 0.798 |
| **CBD-L** | -0.23 | 0.057 | 0.279 | | **CBD-L** | 0.02 | 0.857 | 0.942 |
| **CBD-R** | -0.12 | 0.324 | 0.601 | | **CBD-R** | 0.06 | 0.617 | 0.844 |
| **CBP-L** | -0.07 | 0.544 | 0.798 | | **CBP-L** | 0.02 | 0.881 | 0.946 |
| **CBP-R** | -0.23 | 0.054 | 0.279 | | **CBP-R** | -0.15 | 0.226 | 0.511 |
| **CBT-L** | -0.32 | 0.009 | 0.181 | | **CBT-L** | -0.22 | 0.073 | 0.301 |
| **CBT-R** | -0.13 | 0.291 | 0.554 | | **CBT-R** | -0.14 | 0.250 | 0.521 |
| **CST-L** | 0.19 | 0.115 | 0.373 | | **CST-L** | 0.17 | 0.158 | 0.455 |
| **CST-R** | 0.07 | 0.573 | 0.798 | | **CST-R** | 0.01 | 0.919 | 0.946 |
| **FA-L** | -0.23 | 0.062 | 0.286 | | **FA-L** | -0.08 | 0.500 | 0.796 |
| **FA-R** | -0.02 | 0.841 | 0.937 | | **FA-R** | 0.09 | 0.441 | 0.732 |
| **ILF-L** | -0.01 | 0.951 | 0.963 | | **ILF-L** | 0.17 | 0.173 | 0.472 |
| **ILF-R** | -0.15 | 0.213 | 0.511 | | **ILF-R** | -0.03 | 0.811 | 0.937 |
| **IFO-L** | -0.27 | 0.026 | 0.279 | | **IFO-L** | -0.03 | 0.827 | 0.937 |
| **IFO-R** | -0.37 | 0.003 | 0.166 | | **IFO-R** | -0.14 | 0.265 | 0.529 |
| **MDLF-L** | 0.03 | 0.818 | 0.937 | | **MDLF-L** | 0.14 | 0.254 | 0.521 |
| **MDLF-R** | -0.17 | 0.154 | 0.455 | | **MDLF-R** | -0.14 | 0.236 | 0.511 |
| **OR-L** | -0.25 | 0.040 | 0.279 | | **OR-L** | -0.15 | 0.231 | 0.511 |
| **OR-R** | -0.25 | 0.043 | 0.279 | | **OR-R** | -0.21 | 0.087 | 0.329 |
| **STR-L** | 0.16 | 0.198 | 0.511 | | **STR-L** | 0.13 | 0.288 | 0.554 |
| **STR-R** | 0.02 | 0.841 | 0.937 | | **STR-R** | 0.04 | 0.716 | 0.936 |
| **SLF1-L** | 0.03 | 0.809 | 0.937 | | **SLF1-L** | -0.10 | 0.411 | 0.697 |
| **SLF1-R** | -0.04 | 0.720 | 0.936 | | **SLF1-R** | 0.22 | 0.073 | 0.301 |
| **SLF2-L** | -0.15 | 0.225 | 0.511 | | **SLF2-L** | -0.03 | 0.827 | 0.937 |
| **SLF2-R** | -0.27 | 0.025 | 0.279 | | **SLF2-R** | -0.05 | 0.663 | 0.892 |
| **SLF3-L** | -0.19 | 0.109 | 0.370 | | **SLF3-L** | 0.01 | 0.922 | 0.946 |
| **SLF3-R** | -0.23 | 0.054 | 0.279 | | **SLF3-R** | -0.08 | 0.488 | 0.793 |
| **UF-L** | -0.16 | 0.175 | 0.472 | | **UF-L** | 0.07 | 0.572 | 0.798 |
| **UF-R** | -0.07 | 0.571 | 0.798 | | **UF-R** | 0.03 | 0.805 | 0.937 |
| **VOF-L** | -0.11 | 0.354 | 0.627 | | **VOF-L** | 0.03 | 0.801 | 0.937 |
| **VOF-R** | -0.27 | 0.029 | 0.279 | | **VOF-R** | -0.11 | 0.349 | 0.627 |
| **FMA** | -0.32 | 0.009 | 0.181 | | **FMA** | -0.15 | 0.226 | 0.511 |
| **FMI** | -0.25 | 0.037 | 0.279 | | **FMI** | 0.00 | 0.980 | 0.980 |
| **MCP** | -0.01 | 0.908 | 0.946 | | **MCP** | 0.07 | 0.538 | 0.798 |

**Supplementary Table 4.** Cohen's d effect sizes and raw and adjusted p-values for the comparison of patients with (AH+) and without (AH-) current auditory hallucinations with controls fractional anisotropy (FA).

| | Current AH- | | | | | Current AH+ | | |
|---|---|---|---|---|---|---|---|---|
| **Fibre tract** | **Cohen's d** | **p-value** | **p-value (FDR)** | | **Fibre tract** | **Cohen's d** | **p-value** | **p-value (FDR)** |
| **AF-L** | 0.45 | 0.000 | 0.002 | | **AF-L** | 0.18 | 0.143 | 0.219 |
| **AF-R** | 0.39 | 0.001 | 0.009 | | **AF-R** | 0.21 | 0.090 | 0.163 |
| **AR-L** | 0.26 | 0.030 | 0.064 | | **AR-L** | 0.03 | 0.781 | 0.823 |
| **AR-R** | 0.29 | 0.016 | 0.045 | | **AR-R** | 0.02 | 0.844 | 0.878 |
| **ATR-L** | 0.29 | 0.017 | 0.045 | | **ATR-L** | 0.04 | 0.775 | 0.823 |
| **ATR-R** | 0.19 | 0.121 | 0.201 | | **ATR-R** | 0.06 | 0.617 | 0.707 |
| **CBD-L** | 0.28 | 0.022 | 0.051 | | **CBD-L** | 0.02 | 0.892 | 0.903 |
| **CBD-R** | 0.31 | 0.011 | 0.035 | | **CBD-R** | 0.08 | 0.511 | 0.613 |
| **CBP-L** | 0.26 | 0.034 | 0.069 | | **CBP-L** | 0.01 | 0.905 | 0.905 |
| **CBP-R** | 0.31 | 0.010 | 0.035 | | **CBP-R** | 0.02 | 0.870 | 0.893 |
| **CBT-L** | 0.33 | 0.008 | 0.031 | | **CBT-L** | 0.10 | 0.408 | 0.522 |
| **CBT-R** | 0.24 | 0.044 | 0.089 | | **CBT-R** | 0.04 | 0.726 | 0.786 |
| **CST-L** | 0.29 | 0.016 | 0.045 | | **CST-L** | 0.05 | 0.661 | 0.747 |
| **CST-R** | 0.34 | 0.006 | 0.028 | | **CST-R** | 0.21 | 0.080 | 0.148 |
| **FA-L** | 0.31 | 0.012 | 0.036 | | **FA-L** | 0.14 | 0.243 | 0.344 |
| **FA-R** | 0.20 | 0.104 | 0.180 | | **FA-R** | 0.09 | 0.475 | 0.579 |
| **ILF-L** | 0.46 | 0.000 | 0.002 | | **ILF-L** | 0.19 | 0.117 | 0.199 |
| **ILF-R** | 0.43 | 0.000 | 0.004 | | **ILF-R** | 0.20 | 0.099 | 0.176 |
| **IFO-L** | 0.50 | 0.000 | 0.001 | | **IFO-L** | 0.15 | 0.226 | 0.327 |
| **IFO-R** | 0.46 | 0.000 | 0.002 | | **IFO-R** | 0.23 | 0.062 | 0.118 |
| **MDLF-L** | 0.47 | 0.000 | 0.002 | | **MDLF-L** | 0.18 | 0.140 | 0.219 |
| **MDLF-R** | 0.47 | 0.000 | 0.002 | | **MDLF-R** | 0.29 | 0.017 | 0.045 |
| **OR-L** | 0.50 | 0.000 | 0.001 | | **OR-L** | 0.18 | 0.148 | 0.222 |
| **OR-R** | 0.53 | 0.000 | 0.001 | | **OR-R** | 0.32 | 0.008 | 0.031 |
| **STR-L** | 0.31 | 0.011 | 0.035 | | **STR-L** | 0.06 | 0.616 | 0.707 |
| **STR-R** | 0.29 | 0.019 | 0.045 | | **STR-R** | 0.15 | 0.221 | 0.325 |
| **SLF1-L** | 0.26 | 0.032 | 0.068 | | **SLF1-L** | 0.09 | 0.454 | 0.562 |
| **SLF1-R** | 0.28 | 0.020 | 0.047 | | **SLF1-R** | 0.08 | 0.519 | 0.614 |
| **SLF2-L** | 0.31 | 0.012 | 0.036 | | **SLF2-L** | 0.05 | 0.701 | 0.770 |
| **SLF2-R** | 0.32 | 0.009 | 0.035 | | **SLF2-R** | 0.23 | 0.062 | 0.118 |
| **SLF3-L** | 0.39 | 0.001 | 0.010 | | **SLF3-L** | 0.18 | 0.135 | 0.215 |
| **SLF3-R** | 0.37 | 0.003 | 0.016 | | **SLF3-R** | 0.19 | 0.124 | 0.201 |
| **UF-L** | 0.33 | 0.007 | 0.029 | | **UF-L** | 0.09 | 0.448 | 0.562 |
| **UF-R** | 0.29 | 0.018 | 0.045 | | **UF-R** | 0.13 | 0.295 | 0.403 |
| **VOF-L** | 0.33 | 0.007 | 0.029 | | **VOF-L** | 0.10 | 0.403 | 0.522 |
| **VOF-R** | 0.36 | 0.003 | 0.017 | | **VOF-R** | 0.13 | 0.279 | 0.388 |
| **FMA** | 0.50 | 0.000 | 0.001 | | **FMA** | 0.26 | 0.030 | 0.064 |
| **FMI** | 0.35 | 0.005 | 0.024 | | **FMI** | 0.12 | 0.322 | 0.425 |
| **MCP** | 0.05 | 0.673 | 0.749 | | **MCP** | 0.12 | 0.308 | 0.415 |

**Supplementary Table 5.** Cohen's d effect sizes and raw and adjusted corrected p-values for the comparison of patients with (AH+) and without (AH-) current auditory hallucinations with controls for mean diffusivity (MD).

| | Current AH- | | | | | Current AH+ | | |
|---|---|---|---|---|---|---|---|---|
| **Fibre tract** | **Cohen's d** | **p-value** | **p-value (FDR)** | | **Fibre tract** | **Cohen's d** | **p-value** | **p-value (FDR)** |
| **AF-L** | 0.38 | 0.002 | 0.019 | | **AF-L** | 0.12 | 0.304 | 0.472 |
| **AF-R** | 0.36 | 0.003 | 0.021 | | **AF-R** | 0.17 | 0.155 | 0.301 |
| **AR-L** | 0.07 | 0.541 | 0.660 | | **AR-L** | -0.11 | 0.348 | 0.503 |
| **AR-R** | 0.23 | 0.064 | 0.150 | | **AR-R** | -0.10 | 0.410 | 0.542 |
| **ATR-L** | 0.35 | 0.004 | 0.021 | | **ATR-L** | 0.06 | 0.597 | 0.695 |
| **ATR-R** | 0.23 | 0.056 | 0.140 | | **ATR-R** | 0.11 | 0.363 | 0.515 |
| **CBD-L** | 0.30 | 0.014 | 0.055 | | **CBD-L** | 0.04 | 0.772 | 0.843 |
| **CBD-R** | 0.29 | 0.018 | 0.063 | | **CBD-R** | 0.07 | 0.554 | 0.665 |
| **CBP-L** | 0.16 | 0.179 | 0.333 | | **CBP-L** | -0.03 | 0.808 | 0.852 |
| **CBP-R** | 0.31 | 0.011 | 0.046 | | **CBP-R** | 0.09 | 0.478 | 0.602 |
| **CBT-L** | 0.36 | 0.004 | 0.021 | | **CBT-L** | 0.15 | 0.232 | 0.411 |
| **CBT-R** | 0.22 | 0.066 | 0.150 | | **CBT-R** | 0.09 | 0.479 | 0.602 |
| **CST-L** | 0.10 | 0.425 | 0.553 | | **CST-L** | 0.01 | 0.960 | 0.960 |
| **CST-R** | 0.15 | 0.207 | 0.375 | | **CST-R** | 0.13 | 0.281 | 0.456 |
| **FA-L** | 0.28 | 0.020 | 0.064 | | **FA-L** | 0.12 | 0.334 | 0.492 |
| **FA-R** | 0.16 | 0.179 | 0.333 | | **FA-R** | 0.03 | 0.799 | 0.852 |
| **ILF-L** | 0.30 | 0.015 | 0.055 | | **ILF-L** | 0.06 | 0.638 | 0.732 |
| **ILF-R** | 0.31 | 0.010 | 0.044 | | **ILF-R** | 0.14 | 0.254 | 0.431 |
| **IFO-L** | 0.48 | 0.000 | 0.002 | | **IFO-L** | 0.13 | 0.272 | 0.452 |
| **IFO-R** | 0.47 | 0.000 | 0.002 | | **IFO-R** | 0.24 | 0.050 | 0.130 |
| **MDLF-L** | 0.35 | 0.004 | 0.021 | | **MDLF-L** | 0.12 | 0.315 | 0.472 |
| **MDLF-R** | 0.40 | 0.001 | 0.016 | | **MDLF-R** | 0.28 | 0.024 | 0.071 |
| **OR-L** | 0.49 | 0.000 | 0.002 | | **OR-L** | 0.22 | 0.065 | 0.150 |
| **OR-R** | 0.50 | 0.000 | 0.002 | | **OR-R** | 0.33 | 0.006 | 0.029 |
| **STR-L** | 0.14 | 0.250 | 0.431 | | **STR-L** | -0.03 | 0.824 | 0.857 |
| **STR-R** | 0.21 | 0.088 | 0.190 | | **STR-R** | 0.11 | 0.383 | 0.524 |
| **SLF1-L** | 0.19 | 0.112 | 0.229 | | **SLF1-L** | 0.11 | 0.378 | 0.524 |
| **SLF1-R** | 0.24 | 0.046 | 0.124 | | **SLF1-R** | 0.01 | 0.909 | 0.921 |
| **SLF2-L** | 0.28 | 0.022 | 0.068 | | **SLF2-L** | 0.05 | 0.665 | 0.752 |
| **SLF2-R** | 0.33 | 0.006 | 0.029 | | **SLF2-R** | 0.18 | 0.130 | 0.260 |
| **SLF3-L** | 0.38 | 0.002 | 0.021 | | **SLF3-L** | 0.13 | 0.297 | 0.472 |
| **SLF3-R** | 0.36 | 0.003 | 0.021 | | **SLF3-R** | 0.20 | 0.107 | 0.226 |
| **UF-L** | 0.29 | 0.017 | 0.059 | | **UF-L** | 0.04 | 0.762 | 0.843 |
| **UF-R** | 0.22 | 0.071 | 0.158 | | **UF-R** | 0.08 | 0.531 | 0.657 |
| **VOF-L** | 0.24 | 0.045 | 0.124 | | **VOF-L** | 0.02 | 0.850 | 0.873 |
| **VOF-R** | 0.35 | 0.004 | 0.021 | | **VOF-R** | 0.12 | 0.309 | 0.472 |
| **FMA** | 0.49 | 0.000 | 0.002 | | **FMA** | 0.25 | 0.039 | 0.114 |
| **FMI** | 0.34 | 0.005 | 0.026 | | **FMI** | 0.10 | 0.394 | 0.530 |
| **MCP** | 0.03 | 0.778 | 0.843 | | **MCP** | 0.07 | 0.573 | 0.677 |

**Supplementary Table 6.** Cohen's d effect sizes and raw and adjusted corrected p-values for the comparison of patients with (AH+) and without (AH-) current auditory hallucinations with controls for radial diffusivity (RD).

| | Current AH- | | | | | Current AH+ | | |
|---|---|---|---|---|---|---|---|---|
| Fibre tract | Cohen's d | p-value | p-value (FDR) | | Fibre tract | Cohen's d | p-value | p-value (FDR) |
| AF-L | 0.36 | 0.003 | 0.050 | | AF-L | 0.21 | 0.085 | 0.180 |
| AF-R | 0.35 | 0.004 | 0.050 | | AF-R | 0.20 | 0.105 | 0.201 |
| AR-L | 0.30 | 0.012 | 0.069 | | AR-L | 0.28 | 0.020 | 0.085 |
| AR-R | 0.18 | 0.132 | 0.229 | | AR-R | 0.22 | 0.072 | 0.171 |
| ATR-L | 0.20 | 0.092 | 0.185 | | ATR-L | 0.06 | 0.618 | 0.753 |
| ATR-R | 0.16 | 0.184 | 0.271 | | ATR-R | 0.11 | 0.380 | 0.494 |
| CBD-L | 0.13 | 0.273 | 0.374 | | CBD-L | 0.05 | 0.678 | 0.789 |
| CBD-R | 0.25 | 0.040 | 0.117 | | CBD-R | 0.06 | 0.643 | 0.760 |
| CBP-L | 0.21 | 0.085 | 0.180 | | CBP-L | 0.13 | 0.268 | 0.373 |
| CBP-R | 0.19 | 0.126 | 0.229 | | CBP-R | -0.05 | 0.711 | 0.803 |
| CBT-L | 0.28 | 0.021 | 0.085 | | CBT-L | 0.07 | 0.552 | 0.683 |
| CBT-R | 0.31 | 0.012 | 0.069 | | CBT-R | 0.11 | 0.377 | 0.494 |
| CST-L | 0.28 | 0.023 | 0.085 | | CST-L | 0.02 | 0.865 | 0.912 |
| CST-R | 0.34 | 0.005 | 0.050 | | CST-R | 0.21 | 0.080 | 0.178 |
| FA-L | 0.26 | 0.030 | 0.099 | | FA-L | 0.29 | 0.016 | 0.076 |
| FA-R | 0.20 | 0.107 | 0.201 | | FA-R | 0.18 | 0.129 | 0.229 |
| ILF-L | 0.41 | 0.001 | 0.050 | | ILF-L | 0.31 | 0.010 | 0.069 |
| ILF-R | 0.37 | 0.002 | 0.050 | | ILF-R | 0.23 | 0.059 | 0.154 |
| IFO-L | 0.37 | 0.002 | 0.050 | | IFO-L | 0.10 | 0.411 | 0.526 |
| IFO-R | 0.31 | 0.012 | 0.069 | | IFO-R | 0.18 | 0.146 | 0.242 |
| MDLF-L | 0.34 | 0.005 | 0.050 | | MDLF-L | 0.16 | 0.197 | 0.285 |
| MDLF-R | 0.34 | 0.005 | 0.050 | | MDLF-R | 0.21 | 0.090 | 0.184 |
| OR-L | 0.26 | 0.032 | 0.101 | | OR-L | -0.01 | 0.914 | 0.938 |
| OR-R | 0.31 | 0.011 | 0.069 | | OR-R | 0.18 | 0.150 | 0.243 |
| STR-L | 0.24 | 0.052 | 0.141 | | STR-L | 0.01 | 0.955 | 0.968 |
| STR-R | 0.17 | 0.168 | 0.252 | | STR-R | 0.02 | 0.844 | 0.902 |
| SLF1-L | 0.00 | 0.972 | 0.972 | | SLF1-L | -0.06 | 0.637 | 0.760 |
| SLF1-R | 0.13 | 0.300 | 0.403 | | SLF1-R | -0.01 | 0.908 | 0.938 |
| SLF2-L | 0.17 | 0.161 | 0.251 | | SLF2-L | 0.05 | 0.694 | 0.796 |
| SLF2-R | 0.20 | 0.108 | 0.201 | | SLF2-R | 0.21 | 0.077 | 0.177 |
| SLF3-L | 0.33 | 0.007 | 0.057 | | SLF3-L | 0.28 | 0.023 | 0.085 |
| SLF3-R | 0.27 | 0.025 | 0.089 | | SLF3-R | 0.17 | 0.164 | 0.251 |
| UF-L | 0.30 | 0.015 | 0.076 | | UF-L | 0.22 | 0.068 | 0.166 |
| UF-R | 0.25 | 0.042 | 0.118 | | UF-R | 0.25 | 0.038 | 0.114 |
| VOF-L | 0.22 | 0.066 | 0.166 | | VOF-L | 0.08 | 0.520 | 0.654 |
| VOF-R | 0.18 | 0.145 | 0.242 | | VOF-R | 0.04 | 0.761 | 0.848 |
| FMA | 0.17 | 0.157 | 0.250 | | FMA | 0.03 | 0.831 | 0.900 |
| FMI | 0.27 | 0.029 | 0.099 | | FMI | 0.15 | 0.219 | 0.310 |
| MCP | 0.03 | 0.815 | 0.896 | | MCP | 0.28 | 0.022 | 0.085 |

**Supplementary Table 7.** Cohen's d effect sizes and raw and adjusted corrected p-values for the comparison of patients with (AH+) and without (AH-) current auditory hallucinations with controls for axial diffusivity (AD).

| | SCZ vs CTR | | |
|---|---|---|---|
| Fibre tract | Cohen's d | p-value | p-value (FDR) |
| AF-L | -0.10 | 0.396 | 0.618 |
| AF-R | -0.14 | 0.241 | 0.448 |
| AR-L | 0.32 | 0.008 | 0.111 |
| AR-R | 0.08 | 0.508 | 0.683 |
| ATR-L | -0.29 | 0.015 | 0.121 |
| ATR-R | -0.18 | 0.135 | 0.330 |
| CBD-L | -0.14 | 0.236 | 0.448 |
| CBD-R | -0.05 | 0.694 | 0.804 |
| CBP-L | -0.04 | 0.742 | 0.804 |
| CBP-R | -0.24 | 0.049 | 0.213 |
| CBT-L | -0.33 | 0.006 | 0.111 |
| CBT-R | -0.16 | 0.179 | 0.367 |
| CST-L | 0.22 | 0.066 | 0.256 |
| CST-R | 0.05 | 0.662 | 0.804 |
| FA-L | -0.20 | 0.103 | 0.277 |
| FA-R | 0.04 | 0.774 | 0.804 |
| ILF-L | 0.09 | 0.481 | 0.670 |
| ILF-R | -0.12 | 0.330 | 0.560 |
| IFO-L | -0.20 | 0.104 | 0.277 |
| IFO-R | -0.32 | 0.009 | 0.111 |
| MDLF-L | 0.09 | 0.435 | 0.652 |
| MDLF-R | -0.20 | 0.107 | 0.277 |
| OR-L | -0.25 | 0.041 | 0.213 |
| OR-R | -0.28 | 0.021 | 0.138 |
| STR-L | 0.18 | 0.146 | 0.335 |
| STR-R | 0.04 | 0.737 | 0.804 |
| SLF1-L | -0.04 | 0.774 | 0.804 |
| SLF1-R | 0.09 | 0.461 | 0.665 |
| SLF2-L | -0.11 | 0.347 | 0.564 |
| SLF2-R | -0.21 | 0.079 | 0.277 |
| SLF3-L | -0.12 | 0.303 | 0.538 |
| SLF3-R | -0.20 | 0.092 | 0.277 |
| UF-L | -0.07 | 0.545 | 0.708 |
| UF-R | -0.03 | 0.804 | 0.804 |
| VOF-L | -0.06 | 0.624 | 0.786 |
| VOF-R | -0.24 | 0.047 | 0.213 |
| FMA | -0.30 | 0.015 | 0.121 |
| FMI | -0.17 | 0.155 | 0.336 |
| MCP | 0.03 | 0.796 | 0.804 |

**Supplementary Table 8.** Cohen's d effect sizes and raw and adjusted corrected p-values for the comparison between patients (SCZ) and controls (CTR) for fractional anisotropy (FA).

| | SCZ vs CTR | | |
|---|---|---|---|
| **Fibre tract** | **Cohen's d** | **p-value** | **p-value (FDR)** |
| **AF-L** | 0.40 | 0.001 | 0.004 |
| **AF-R** | 0.38 | 0.002 | 0.007 |
| **AR-L** | 0.20 | 0.105 | 0.121 |
| **AR-R** | 0.21 | 0.084 | 0.103 |
| **ATR-L** | 0.21 | 0.078 | 0.098 |
| **ATR-R** | 0.16 | 0.186 | 0.190 |
| **CBD-L** | 0.20 | 0.105 | 0.121 |
| **CBD-R** | 0.25 | 0.037 | 0.061 |
| **CBP-L** | 0.18 | 0.134 | 0.143 |
| **CBP-R** | 0.22 | 0.069 | 0.090 |
| **CBT-L** | 0.27 | 0.024 | 0.044 |
| **CBT-R** | 0.19 | 0.121 | 0.135 |
| **CST-L** | 0.23 | 0.062 | 0.085 |
| **CST-R** | 0.34 | 0.005 | 0.013 |
| **FA-L** | 0.28 | 0.019 | 0.042 |
| **FA-R** | 0.18 | 0.136 | 0.143 |
| **ILF-L** | 0.41 | 0.001 | 0.004 |
| **ILF-R** | 0.40 | 0.001 | 0.004 |
| **IFO-L** | 0.41 | 0.001 | 0.004 |
| **IFO-R** | 0.44 | 0.000 | 0.003 |
| **MDLF-L** | 0.41 | 0.001 | 0.004 |
| **MDLF-R** | 0.48 | 0.000 | 0.001 |
| **OR-L** | 0.43 | 0.000 | 0.003 |
| **OR-R** | 0.53 | 0.000 | 0.001 |
| **STR-L** | 0.24 | 0.046 | 0.072 |
| **STR-R** | 0.27 | 0.024 | 0.044 |
| **SLF1-L** | 0.23 | 0.063 | 0.085 |
| **SLF1-R** | 0.23 | 0.054 | 0.080 |
| **SLF2-L** | 0.23 | 0.056 | 0.080 |
| **SLF2-R** | 0.34 | 0.006 | 0.014 |
| **SLF3-L** | 0.36 | 0.003 | 0.010 |
| **SLF3-R** | 0.35 | 0.004 | 0.012 |
| **UF-L** | 0.27 | 0.025 | 0.044 |
| **UF-R** | 0.26 | 0.030 | 0.050 |
| **VOF-L** | 0.28 | 0.022 | 0.044 |
| **VOF-R** | 0.32 | 0.010 | 0.023 |
| **FMA** | 0.48 | 0.000 | 0.001 |
| **FMI** | 0.30 | 0.014 | 0.033 |
| **MCP** | 0.10 | 0.398 | 0.398 |

**Supplementary Table 9.** Cohen's d effect sizes and raw and adjusted corrected p-values for the comparison between patients (SCZ) and controls (CTR) for mean diffusivity (MD).

| | SCZ vs CTR | | |
|---|---|---|---|
| Fibre tract | Cohen's d | p-value | p-value (FDR) |
| AF-L | 0.33 | 0.007 | 0.029 |
| AF-R | 0.34 | 0.006 | 0.028 |
| AR-L | -0.01 | 0.919 | 0.919 |
| AR-R | 0.10 | 0.424 | 0.487 |
| ATR-L | 0.27 | 0.025 | 0.058 |
| ATR-R | 0.22 | 0.073 | 0.117 |
| CBD-L | 0.22 | 0.070 | 0.117 |
| CBD-R | 0.23 | 0.056 | 0.107 |
| CBP-L | 0.09 | 0.439 | 0.489 |
| CBP-R | 0.25 | 0.036 | 0.073 |
| CBT-L | 0.32 | 0.009 | 0.029 |
| CBT-R | 0.20 | 0.102 | 0.148 |
| CST-L | 0.07 | 0.571 | 0.601 |
| CST-R | 0.18 | 0.148 | 0.187 |
| FA-L | 0.26 | 0.035 | 0.073 |
| FA-R | 0.13 | 0.294 | 0.347 |
| ILF-L | 0.23 | 0.057 | 0.107 |
| ILF-R | 0.29 | 0.018 | 0.045 |
| IFO-L | 0.40 | 0.001 | 0.008 |
| IFO-R | 0.45 | 0.000 | 0.002 |
| MDLF-L | 0.30 | 0.012 | 0.034 |
| MDLF-R | 0.42 | 0.001 | 0.005 |
| OR-L | 0.45 | 0.000 | 0.002 |
| OR-R | 0.52 | 0.000 | 0.001 |
| STR-L | 0.08 | 0.512 | 0.555 |
| STR-R | 0.20 | 0.102 | 0.148 |
| SLF1-L | 0.19 | 0.119 | 0.160 |
| SLF1-R | 0.17 | 0.159 | 0.194 |
| SLF2-L | 0.22 | 0.074 | 0.117 |
| SLF2-R | 0.33 | 0.008 | 0.029 |
| SLF3-L | 0.32 | 0.008 | 0.029 |
| SLF3-R | 0.35 | 0.004 | 0.023 |
| UF-L | 0.22 | 0.075 | 0.117 |
| UF-R | 0.19 | 0.118 | 0.160 |
| VOF-L | 0.18 | 0.146 | 0.187 |
| VOF-R | 0.30 | 0.012 | 0.034 |
| FMA | 0.47 | 0.000 | 0.002 |
| FMI | 0.29 | 0.018 | 0.045 |
| MCP | 0.06 | 0.617 | 0.633 |

**Supplementary Table 10.** Cohen's d effect sizes and raw and adjusted corrected p-values for the comparison between patients (SCZ) and controls (CTR) for radial diffusivity (RD).

| | SCZ vs CTR | | |
|---|---|---|---|
| Fibre tract | Cohen's d | p-value | p-value (FDR) |
| AF-L | 0.36 | 0.003 | 0.023 |
| AF-R | 0.34 | 0.005 | 0.023 |
| AR-L | 0.36 | 0.003 | 0.023 |
| AR-R | 0.24 | 0.045 | 0.088 |
| ATR-L | 0.17 | 0.158 | 0.218 |
| ATR-R | 0.17 | 0.168 | 0.218 |
| CBD-L | 0.12 | 0.332 | 0.360 |
| CBD-R | 0.20 | 0.101 | 0.160 |
| CBP-L | 0.21 | 0.077 | 0.130 |
| CBP-R | 0.10 | 0.407 | 0.429 |
| CBT-L | 0.23 | 0.059 | 0.105 |
| CBT-R | 0.27 | 0.029 | 0.066 |
| CST-L | 0.20 | 0.103 | 0.160 |
| CST-R | 0.35 | 0.004 | 0.023 |
| FA-L | 0.34 | 0.005 | 0.023 |
| FA-R | 0.23 | 0.055 | 0.102 |
| ILF-L | 0.45 | 0.000 | 0.008 |
| ILF-R | 0.38 | 0.002 | 0.023 |
| IFO-L | 0.30 | 0.012 | 0.033 |
| IFO-R | 0.30 | 0.013 | 0.033 |
| MDLF-L | 0.32 | 0.009 | 0.033 |
| MDLF-R | 0.34 | 0.005 | 0.023 |
| OR-L | 0.17 | 0.166 | 0.218 |
| OR-R | 0.30 | 0.012 | 0.033 |
| STR-L | 0.16 | 0.180 | 0.226 |
| STR-R | 0.13 | 0.298 | 0.332 |
| SLF1-L | -0.03 | 0.777 | 0.777 |
| SLF1-R | 0.08 | 0.524 | 0.538 |
| SLF2-L | 0.14 | 0.244 | 0.294 |
| SLF2-R | 0.25 | 0.040 | 0.082 |
| SLF3-L | 0.38 | 0.002 | 0.023 |
| SLF3-R | 0.28 | 0.023 | 0.055 |
| UF-L | 0.32 | 0.008 | 0.032 |
| UF-R | 0.31 | 0.012 | 0.033 |
| VOF-L | 0.19 | 0.111 | 0.166 |
| VOF-R | 0.14 | 0.249 | 0.294 |
| FMA | 0.13 | 0.283 | 0.324 |
| FMI | 0.26 | 0.032 | 0.069 |
| MCP | 0.17 | 0.160 | 0.218 |

**Supplementary Table 11.** Cohen's d effect sizes and raw and adjusted corrected p-values for the comparison between patients (SCZ) and controls (CTR) for axial diffusivity (AD).

| Fibre tract | FA | MD | RD | AD |
|---|---|---|---|---|
| AF-L | -0.79 | 0.93 | 0.91 | 0.87 |
| AF-R | -0.81 | 0.92 | 0.90 | 0.89 |
| AR-L | -0.43 | 0.74 | 0.63 | 0.56 |
| AR-R | -0.32 | 0.69 | 0.51 | 0.60 |
| ATR-L | -0.74 | 0.84 | 0.81 | 0.82 |
| ATR-R | -0.72 | 0.83 | 0.80 | 0.80 |
| CBD-L | -0.63 | 0.80 | 0.76 | 0.64 |
| CBD-R | -0.58 | 0.76 | 0.71 | 0.67 |
| CBP-L | -0.50 | 0.61 | 0.59 | 0.42 |
| CBP-R | -0.38 | 0.59 | 0.54 | 0.48 |
| CBT-L | -0.39 | 0.58 | 0.50 | 0.48 |
| CBT-R | -0.32 | 0.48 | 0.41 | 0.42 |
| CST-L | -0.57 | 0.82 | 0.74 | 0.64 |
| CST-R | -0.59 | 0.84 | 0.75 | 0.68 |
| FA-L | -0.68 | 0.79 | 0.77 | 0.73 |
| FA-R | -0.60 | 0.76 | 0.72 | 0.74 |
| ILF-L | -0.63 | 0.87 | 0.81 | 0.74 |
| ILF-R | -0.65 | 0.88 | 0.80 | 0.78 |
| IFO-L | -0.88 | 0.94 | 0.93 | 0.88 |
| IFO-R | -0.88 | 0.93 | 0.92 | 0.89 |
| MDLF-L | -0.77 | 0.92 | 0.90 | 0.85 |
| MDLF-R | -0.79 | 0.91 | 0.89 | 0.82 |
| OR-L | -0.77 | 0.87 | 0.86 | 0.77 |
| OR-R | -0.80 | 0.87 | 0.86 | 0.77 |
| STR-L | -0.65 | 0.86 | 0.83 | 0.69 |
| STR-R | -0.61 | 0.84 | 0.79 | 0.73 |
| SLF1-L | -0.68 | 0.85 | 0.82 | 0.72 |
| SLF1-R | -0.62 | 0.82 | 0.78 | 0.72 |
| SLF2-L | -0.49 | 0.80 | 0.72 | 0.80 |
| SLF2-R | -0.51 | 0.78 | 0.70 | 0.81 |
| SLF3-L | -0.68 | 0.89 | 0.85 | 0.84 |
| SLF3-R | -0.69 | 0.86 | 0.83 | 0.83 |
| UF-L | -0.72 | 0.80 | 0.77 | 0.74 |
| UF-R | -0.64 | 0.75 | 0.71 | 0.70 |
| VOF-L | -0.50 | 0.70 | 0.67 | 0.61 |
| VOF-R | -0.49 | 0.73 | 0.68 | 0.63 |
| FMA | -0.66 | 0.77 | 0.75 | 0.68 |
| FMI | -0.79 | 0.83 | 0.83 | 0.76 |
| MCP | -0.35 | 0.33 | 0.32 | 0.33 |

**Supplementary Table 12.** Correlations between tract-wise DTI metrics and unimodal g-factors.

| | Correlation | | Contribution | |
|---|---|---|---|---|
| | **g-Dim1** | **g-Dim2** | **g-Dim1** | **g-Dim2** |
| **FA** | -0.72 | -0.66 | 19.94 | 33.90 |
| **MD** | 0.95 | -0.25 | 35.25 | 4.64 |
| **RD** | 0.99 | 0.13 | 37.82 | 1.40 |
| **AD** | 0.42 | -0.88 | 6.99 | 60.07 |

**Supplementary Table 13.** Correlations (left) and percentage contributions (right) between each DTI metric and the multimodal g-factors, g-Dim1 and g-Dim2.

Preprint version

**Supplementary Note 1.** Clinical assessment of auditory hallucinations

The AH+ group was comprised of 49 patients with schizophrenia (83.1%) and 10 patients with schizoaffective disorder (16.9%), and the AH- group was comprised of 57 patients with schizophrenia (70.4%), 14 patients with schizoaffective disorder (17.3%), and 10 patients with schizophreniform disorder (12.3%).

The lifetime AH+ group was comprised of 69 patients with schizophrenia (74.2%) and 20 patients with schizoaffective disorder (21.5%) and 4 patients with schizophreniform disorder (4.3%). The lifetime AH- group was comprised of 24 patients with schizophrenia (72.7%), 3 patients with schizoaffective disorder (9.1%), and 6 patients with schizophreniform disorder (12.3%). A total of 14 patients were missing lifetime AH data.

In the TOP dataset, we used the PANSS P3 item to assess current AH status. The median time between assessment with PANSS and MRI acquisition was 12 days. PANSS P3 assesses "hallucinatory behaviour" including hallucinations in auditory, visual, olfactory or somatic modalities. As such, this item is not specific to AH. However, hallucinations in patients with SCZ most commonly present as AH (Goodwin & Rosenthal, 1971; Lim et al., 2016).

Patients in the HUBIN dataset were assessed with the SAPS, which distinguishes between AH and hallucinations in other modalities. The median time between assessment with SAPS and MRI acquisition was 0 days. In this dataset, 96.1% of participants (all but one) with hallucinations in the somatic, olfactory or visual modalities (items H4, H5, and H6 > 1) also had AH. This supported our interpretation that PANSS P3 mostly reflects AH.

Preprint version

**Supplementary Note 2.** Selection of fibre tracts

The XTRACT toolbox implements tractography protocols for reconstructing 42 intra- and interhemispheric fibre tracts. In the present study, some fibre tracts could not be reliably reconstructed for all participants, resulting in empty masks. These were the left and right fornix (FX) and the interhemispheric anterior commissure (AC). To ensure the inclusion of as many patients as possible, we excluded these from the final analyses resulting in the inclusion of 39 fibre tracts. See **Supplementary Table 13** for a list of the included fibre tracts.

Preprint version

**Supplementary Note 3.** Quality assurance of diffusion tensor imaging data

To assess data quality, we created subject-wise image summaries for each DTI metric and FA maps with whole-brain segmentations based on BET (Smith, 2002) and the standard FSL FMRIB58 FA template in MNI space, as well as probabilistic segmentations of fibre tracts. The image summaries were visually inspected for all participants to assess image quality and potential fibre tract reconstruction errors. See **Supplementary Figure 6** for an example image summary for four fibre tracts in a single participant.

As a further quality control of fibre tract reconstruction, we identified outliers using the Median Absolute Deviation (MAD; Leys, Ley, Klein, Bernard, & Licata, 2013) method from the *Routliers* package (https://CRAN.R-project.org/package=Routliers) with a threshold of 5. Outliers were inspected with *xtract_viewer* (https://fsl.fmrib.ox.ac.uk/fsl/fslwiki/XTRACT) and removed from all analyses in the case of major reconstruction errors. Finally, we visually assessed DTI metric distributions to ensure there were no violations of the assumptions of the statistical analyses. See **Supplementary Figure 2-5** for tract-wise histograms for each DTI metric.

As described in **Supplementary Note 2**, some fibre tracts could not be reconstructed reliably. This issue was also present among the 39 fibre selected tracts and we therefore excluded 22 participants (4 patients) due to empty masks. Of these 12 had empty left CBP masks followed by 5 participants with empty MCP masks. We further excluded one participant due to missing DTI data and one participant due to image quality. Four additional participants were flagged as outliers and excluded due to fibre tract reconstruction errors. All quality assurance was performed blinded to the group membership of the participant.

Preprint version

**Supplementary Note 4.** Tract-wise group differences in patients with and without a history of lifetime AH

No significant group differences were observed between patients with (L-AH+) or without (L-AH-) a history of lifetime AH and healthy controls for FA or AD. Higher MD was observed in L-AH- compared to controls, for the AF, FA, ILF, IFO, MDLF, OR, SLF1, SLF2 in the right hemisphere, as well as the bilateral SLF3 and FMI. We also observed higher MD in L-AH+ compared to controls in the left AF, left CBT, left ILF, bilateral IFO, bilateral MDLF, bilateral OR, and FMA.

Higher RD was observed in L-AH- compared to controls in the FA, ILF, right IFO, OR, and SLF3 in the right hemisphere, as well as the FMI. Higher RD was observed in L-AH+ compared to controls in the left CBT, bilateral IFO, right MDLF, bilateral OR, and FMA. There were no significant differences between L-AH+ and L-AH- after correction for multiple testing for any of the DTI metrics.

Preprint version

**Supplementary Note 5.** Tract-wise group differences in patients compared to controls

For comparison with past studies on DTI metrics in patients with SCZ, we examined group differences between patients with SCZ and healthy controls in DTI metrics (FA, RD, MD, AD). We used regression models adjusted for age, $age^2$, and sex, with group (healthy control or patient) as variable of interest.

No significant differences between SCZ and controls were observed for FA. Lower MD was observed in SCZ for the bilateral AF, left CBT, right CST, left FA, bilateral ILF, bilateral IFO, bilateral MDLF, bilateral OR, right STR, right SLF2, bilateral SLF3, left UF, bilateral VOF, FMA, and FMI compared to controls.

Higher RD was observed in SCZ for the bilateral AF, left CBT, right ILF, bilateral IFO, bilateral MDLF, bilateral OR, right SLF2, bilateral SLF3, right VOF, FMA, and FMI compared to controls. Higher AD was observed in SCZ for the bilateral AF, left AR, right CST, left FA, bilateral ILF, bilateral IFO, bilateral MDLF, right OR, left SLF3, and bilateral UF compared to controls.

See **Supplementary Figure 8** and **9** for bar plots of Cohen's d effect sizes and **Supplementary Tables 8-11** for Cohen's d effect sizes and adjusted and unadjusted p-values for each DTI metric.

Preprint version


Preprint version
